# Outcomes and Optimization of Two Reading Training Protocols for Individuals with Homonymous Visual Field Defects

**DOI:** 10.1101/2025.11.03.25339232

**Authors:** S. Tol, J. Heutink, E. J. Veldman, J. Koopman, J. M. Vermeer, V. Buijnink, G.A. de Haan

## Abstract

Individuals with Homonymous Visual Field Defects (HVFDs) after brain injury often experience difficulty in reading. This can be debilitating, as reading is an important aspect of daily life. The current study aims to explore the potential outcomes of two reading trainings for individuals with HVFDs. One training, the Vistra reading training, aims at compensating for the HVFD by practicing adjusted reading eye movements. The second training is the Rotated Reading training, which aims to utilize the intact visual field by practicing to read rotated text in a personalized angle. Next to assessing the potential outcomes of the Vistra and Rotated Reading trainings on a group and individual level, this study aims to investigate patient experiences with the trainings. Using a Partially Randomized Patient Preferred Controlled Trial, participants were assigned to one of three groups: Vistra, Rotated Reading or a waiting list control group. There were 5 participants included in both the Vistra and Rotated Reading groups, and 10 in the control group. Assessments were conducted before(T1), after(T2) and 3 months after(T3) training or the waiting period, including reading speed, self-reported reading efficacy, attitude, skills, and objects, and quality of life. Results showed significant group-level improvements following Vistra training on reading speed, self-reported reading skills and reading objects. Furthermore, within some Vistra participants, significant individual changes were observed on reading speed, self-reported reading efficacy, reading objects and quality of life. No significant group effects were found for Rotated Reading training, but some individuals demonstrated significant individual changes on self-reported reading efficacy and reading skills. Qualitative analysis identified five key themes of reading training outcome: reading objects, reading skills, reading enjoyment, reading certainty and non-reading related outcomes. Participants reported adopting specific reading strategies and emphasized perceived improvement as a key motivator for continued use. Feedback highlighted the wish for personalized training approaches.

## Introduction

Individuals with Homonymous Visual Field Defects (HVFDs) often experience difficulty in reading (1,2). This can be debilitating, as reading is an important aspect of daily life. For instance, reading is positively related to health and cognition and needed for work and social life (3–8). HVFDs are caused by acquired brain injury to post-chiasmal nerves in the brain, causing the individual with HVFDs (iwHs) to experience blindness in one half or quarter of the visual field (2,9). In iwHs, the processes of visual input (bottom-up processing) and attentional control (top-down processing) involved in reading and guiding reading eye movements are disrupted (10). Consequently, iwHs experience difficulties in recognizing words, finding the beginning or end of a line, skipping (parts) of words or erroneous completion of incomplete read words. Due to increased reading errors and inadequate reading eye movements, iwHs often show significantly lower reading speed, compared to individuals without HVFDs (2,10–14).

Current knowledge derived from systematic reviews and meta-analyses indicates that trainings aiming to compensate for the HVFD through compensatory eye movements are most promising to increase reading speed in iwHs (15,16). Recently, two reading trainings for iwHs have been developed in the Netherlands at two centres of expertise for the visually impaired and blind (Royal Dutch Visio and Bartiméus). In the Netherlands, neurorehabilitation at such expertise centres is available for all citizens with a professional referral by a medical specialist (e.g. by a neurologist) under the Health Care Insurance Act. At the expertise centres, care professionals from different disciplines (e.g. occupational therapists, neuropsychologists, vision experts) have specialized in HVFD rehabilitation. These visual neurorehabilitation professionals have developed the two reading trainings, which are already provided to iwHs in Dutch clinical practice at Royal Dutch Visio and Bartiméus. One training, the Vistra reading training, aims at compensating for the HVFD by practicing adjusted eye movements during reading more fitting to the new visual field (e.g. smaller eye jumps or ‘saccades’), as well as other compensatory behaviors to gain better overview of the text. With this compensatory approach, it shows resemblance to earlier studied interventions (13,17–19), which have been shown to increase reading speed (15). The difference between these studies and Vistra is that, whereas these previous studies include single-word reading tasks with increasing length and presentation time, the Vistra also incorporates sentences, paragraphs, psychoeducation and practice with different reading objects. The second training is the Rotated Reading training, which aims to utilize the intact visual field by practicing to read rotated text in a personalized angle. This novel concept of learning to read rotated text in HVFDs has been studied before (11,20,21). These authors all studied horizontal and vertical reading training, and found that this training type has beneficial effects on reading speed in iwHs (however, the effect of horizontal reading training was found to be larger compared to vertical reading training). In the Rotated Reading training studied in the current study, the angle of rotation is personalized for every iwH, dependent on their exact central visual field loss. More information about the protocols of both trainings can be found in the Methods section. The current study is the first to present the Vistra reading training and Rotated Reading training.

The scientifically studied effectiveness of interventions is one source of evidence that should be considered in clinical decision making for individual patients, according to Evidence Based Medicine (EBP) (22,23). Other sources that should aid clinical decision making, according to EBP, are the abilities of the professional and the patient preferences and experiences (22). Such information from the patient’s side could be gained by self-report assessments and qualitative interviews. To assess the effects of HVFD interventions on reading, reading speed (read out loud) has been the most used measure (24). Besides reading speed, other used reading measures are reading errors, self-reported reading difficulties and reading eye-movement parameters. The potential of including self-report measures of multiple aspects of reading, as well as measures that capture individually relevant outcomes has been underexplored in HVFD intervention research (24–26). Our most recent study has shown that iwHs experience a broad range of reading difficulties (27).

The current study aims to contribute to the scientific evidence for the effects of the Vistra and Rotated Reading trainings, and the training experiences of the patients. This is done by focusing on: 1) a data-driven exploration of the potential training outcomes (both on a group level and individual level) taking into account Vistra, Rotated Reading and a control group, and 2) an investigation of patient experiences with the trainings, including outcomes, applicability and feedback. Reading speed and self-reported relationship to reading, reading skills and daily life reading of different objects were taken into account. Furthermore, vision-related quality of life was included. Lastly, semi-structured interviews were conducted with the participants who followed the reading trainings focusing on training outcomes, use of the learned strategies and participants’ feedback on the trainings. The outcomes of this study are relevant for both current EBP decision making and future HVFD studies concerning reading.

## Method

### Trainings

#### Vistra Reading Training

Vistra is a saccadic compensation reading training for iwHs. There are seven themes within the training, which can be covered in approximately 8-10 sessions of 60-90 minutes, one session a week. Vistra is provided by a trained occupational therapist. Every theme takes into account the learned strategies of the previous themes. The general concept of Vistra is to compensate for the visual field loss, including strategies to create overview and adjusted eye movements during reading. The seven themes are: 1) insight and psychoeducation on reading and HVFDs, 2) strategies to gain overview of different reading sources, 3) practice with adjusted eye movements, 4) for right-sided HVFDs: practice with adjusted eye movements during reading/continued reading; for left-sided HVFDs: practice with return sweep, 5) strategies to keep place/line in text, 6) practice with continued reading with even pace and 7) generalization of learned strategies to different reading objects and situations. To practice the adjusted eye movements, a software program is used with a fixation cross and stimuli in the blind hemifield with differing properties and presentation time. The participant starts practicing with single-figure stimuli, which builds up to words, sentences and eventually paragraphs. In between sessions, iwHs are provided with daily homework assignments on paper which should take around 15 minutes to complete. More information on Vistra can be found in the TIDieR checklist for reporting interventions in Supplementary file S1 (28).

#### Rotated Reading Training

The Rotated Reading training is aimed at utilizing the intact central visual field by rotating text into a personalized angle dependent on the visual field loss of the iwH. In finding the right personalized angle, there is focus on the central visual field loss (central 5 degrees), as this covers the region of effective vision for reading in which letters can be identified (i.e. the perceptual span; Rayner, 1998; Schuett et al., 2008b). In iwHs, the degrees of central sparing (also known as macular sparing) can differ regardless of the size of the HVFD, dependent on the exact location of the neural damage causing the HVFDs (e.g. 2). The training includes approximately five weekly sessions of 60 minutes, provided by a trained occupational therapist. Further, the participant is asked to make daily homework assignments of 15 minutes. Every session adresses two sub-goals, leading to a total of 10 sub-goals (see Table 1). The TIDieR checklist for reporting interventions of the Rotated Reading training can be found in Supplemantary file S1 (28).

**Table 1.** Overview of Subgoals per Session of the Rotated Reading Training.

Overview of Subgoals per Session of the Rotated Reading Training
| Session | Subgoals |
| --- | --- |
| 1 | 1) determining the correct angle of rotation |
|  | 2) providing insight and psychoeducation on reading and HVFDs |
| 2 | 3) gaining overview of rotated text |
|  | 4) reading rotated words of maximal 7 letters |
| 3 | 5) reading compound words of maximal 12 letters |
|  | 6) reading rotated sentences |
| 4 | 7) reading short rotated texts |
|  | 8) reading rotated meaningful texts (e.g. personal book of participant) |
| 5 | 9) evaluation of the progress made |
|  | 10) discussion of how to implement the learned strategies in daily life |

### Study Design

The current study started out as a Randomized Controlled Trial (RCT) which was revised during the data collection to a Partially Randomized Patient Preferred Controlled Trial (PRPPCT) in order to facilitate higher inclusion rates. The participants were iwHs who received neurorehabilitation at Visio or Bartiméus. Their decision whether or not to participate in this study did not affect their rehabilitation options. Three groups were compared: Vistra, Rotated Reading training and a waiting list control group (see Figure 1). Participants were assessed at three time points: 0-3 weeks before the training or waiting period (T1), 0-3 weeks after the training or waiting period (T2) and three months after the training (T3). As participants in the waiting list control group had the opportunity to follow reading training after T2, no assessment on T3 was administered in this group. Assessments T1 and T2 were administered face-to-face at the rehabilitation centers, T3 was administered through an online video call. The method for allocating participants to one of the three groups was adaptive randomization, also called minimization. This procedure is based on the idea that with every allocation, the differences between the groups on certain relevant patient characteristics are minimized (31). This randomization method has often been recommended in clinical trials, especially in studies with smaller sample sizes (32). In the current study, the minimization technique was applied to ensure maximal balance in the three study groups regarding (1) side (left vs. right) and (2) type (hemianopia vs. quadrantanopia) of visual field defect, (3) amount of macular sparing (sparing vs. splitting) and (4) organization (Visio vs. Bartiméus). Due to continuing low inclusion rates after five months of the RCT, an amendment was made which included changing the design from a three-group RCT to a PRPPCT, in which participants have the option to choose which study group they would want to enter, if they had a preference. In case of no preference, the investigator allocated the participant according to the minimization method. A recent meta-analysis (33) on the PRPPCT design in health care research has shown that incorporating participant preference into a controlled trial can lead to substantial increase of participants. Based on multiple parameters such as the influence of preference on outcome measures, characteristics of participants with or without preference and dropout rates, the authors concluded that the PRPPCT design has the potential to increase external validity while maintaining internal validity. A second change within the amendment was to expand the inclusion criteria for the waiting list control group to iwHs who will not follow reading training or prioritize another training programme (in practice, often mobility training). Experience from clinical practice also shows that patients who do not (immediately) follow reading training in their rehabilitation programme often do have reading complaints and problems. After ethical approval, the PRPPT was implemented 6 months before the end of the inclusion period.

**Figure 1.**
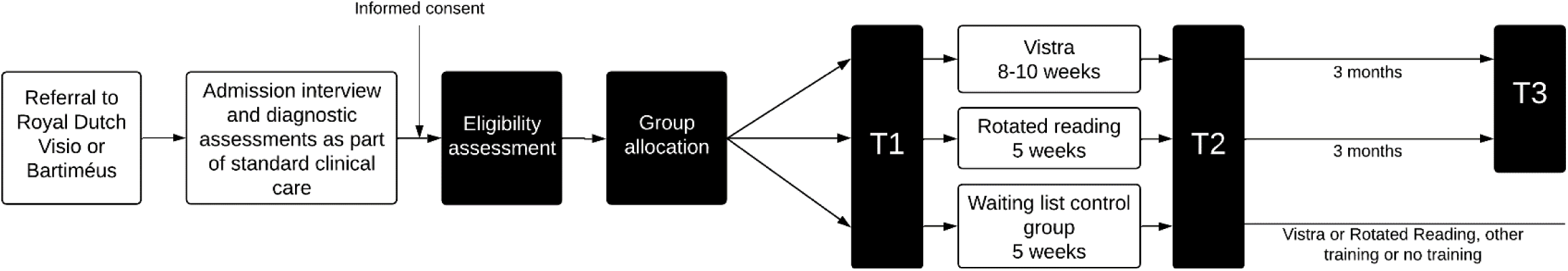
Schematic Overview of Study Design. *Note.* White boxes reflect activities related to standard clinical care, black boxes reflect activities related to research. Assessments for the control group were administered within a regular waiting period standard for every participant in clinical care, meaning that participation in the study did not delay their rehabilitation.

Prior to the RCT data collection, a pilot RCT was conducted in which the RCT structure was implemented together with feasibility measures such as the amount of potential and final inclusions, duration of assessments or protocol adherence. The duration of the pilot phase was six months, after which only one participant could be included due to varying (mostly organizational) reasons. After the pilot phase, adaptations were made to the study protocol to facilitate higher inclusion rates (e.g. removing participation barriers such as potential long waiting time before training and including digital auditory participant information). As these changes did not regard the training protocols and outcome measures, it was decided to include the participant from the pilot phase into the final dataset.

### Ethics and Registration

The protocol of the current study was approved by the Medical Ethics Review Committee of the University Medical Centre Groningen, registration number METc 2021/199 (Pilot) and METc 2022/120 (RCT/PRPPT). The study was performed in accordance with the Declaration of Helsinki. Further, the protocol was registered as a clinical trial at the ISRCTN Registry, ID ISRCTN85915866 (34). All participants gave informed consent.

### Participants

Participants were recruited at Royal Dutch Visio and Bartiméus. Inclusion criteria were: Homonymous visual field defect (at least a quadrantanopia, either right-sided or left-sided) due to acquired post-chiasmatic brain injury, at least three months between onset HVFD and the first measurement at T1, binocular near visual acuity of at least Snellen 0.5 (6/12 or 20/40, LogMAR 0.3) with the individuals own current correction, a Mini-Mental State Examination score of 24 or higher (35,36), age of 18 or older, and the presence of a reading-related treatment goal (except for individuals included in the control group in the PRPPT). Exclusion criteria were: additional visual field defect in ipsilesional visual hemi-field, no clear neurological cause of HVFD, presence of comorbid neglect as indicated by an assessment of the combined scores on the Bells test (37), Line Bisection task (38), Clock drawing task (39) and a spontaneous drawing task. Further, problems leading to inability to follow the reading training were formulated as exclusion criteria, such as illiteracy, low literacy, other pre-morbid reading difficulties, communication difficulties (e.g. severe hearing impairment, no fluent understanding of Dutch language, severe aphasia as indicated by the Token test, part 5) (40), additional visual disturbances (e.g. diplopia, low contrast sensitivity), and negative advice of treatment team regarding reading training participation, due to e.g. severe psychiatric, cognitive or visual perception disorders, problems with health, motivation or illness awareness or substance misuse.

Recruitment of potential participants lasted for 20 months in total. Within this time, 24 participants could be included. Of these 24, four participants were excluded for the current analyses. For two participants this was because only data could be collected at T1. Of these two participants, one participant did not wish to continue with the study after T1, and the second participant suffered a second stroke during training. Two other participants were excluded for analysis due to changes in the visual field loss during training. Of the 20 participants included in the final dataset, five followed Vistra, five Rotated Reading training and 10 participants were included in the waiting list control group. An overview of the participant characteristics can be found in Table 2.

**Table 2.**
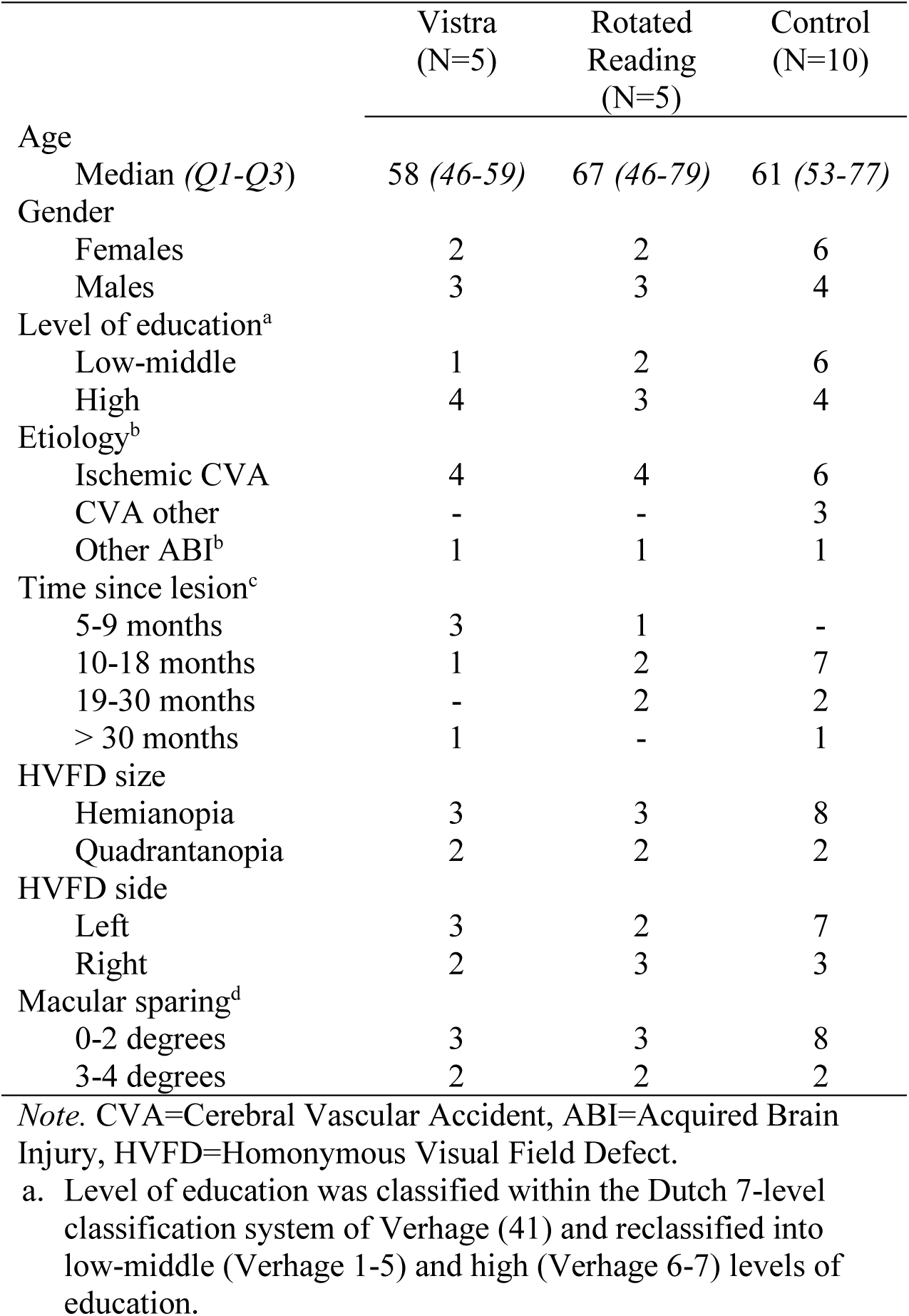
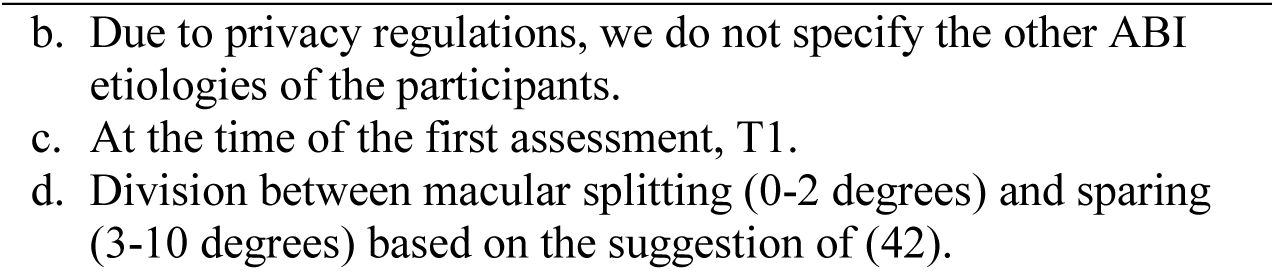
Overview of Demographic and Clinical Characteristics of the Participants.

|  | Vistra<br>(N=5) | Rotated<br>Reading<br>(N=5) | Control<br>(N=10) |
| --- | --- | --- | --- |
| Age |  |  |  |
| Median (*Q1-Q3*) | 58 (46-59) | 67 (46-79) | 61 (53-77) |
| Gender |  |  |  |
| Females | 2 | 2 | 6 |
| Males | 3 | 3 | 4 |
| Level of education<sup>a</sup> |  |  |  |
| Low-middle | 1 | 2 | 6 |
| High | 4 | 3 | 4 |
| Etiology<sup>b</sup> |  |  |  |
| Ischemic CVA | 4 | 4 | 6 |
| CVA other | - | - | 3 |
| Other ABI<sup>b</sup> | 1 | 1 | 1 |
| Time since lesion<sup>c</sup> |  |  |  |
| 5-9 months | 3 | 1 | - |
| 10-18 months | 1 | 2 | 7 |
| 19-30 months | - | 2 | 2 |
| > 30 months | 1 | - | 1 |
| HVFD size |  |  |  |
| Hemianopia | 3 | 3 | 8 |
| Quadrantanopia | 2 | 2 | 2 |
| HVFD side |  |  |  |
| Left | 3 | 2 | 7 |
| Right | 2 | 3 | 3 |
| Macular sparing<sup>d</sup> |  |  |  |
| 0-2 degrees | 3 | 3 | 8 |
| 3-4 degrees | 2 | 2 | 2 |

Note. CVA=Cerebral Vascular Accident, ABI=Acquired Brain Injury, HVFD=Homonymous Visual Field Defect.
- a. Level of education was classified within the Dutch 7-level classification system of Verhage (41) and reclassified into low-middle (Verhage 1-5) and high (Verhage 6-7) levels of education.
- - b. Due to privacy regulations, we do not specify the other ABI etiologies of the participants. - c. At the time of the first assessment, T1. - d. Division between macular splitting (0-2 degrees) and sparing (3-10 degrees) based on the suggestion of (42).

### Materials

#### Quantitative Measures of Reading and Quality of Life

##### Words Per Minute

Objective reading performance was measured with the International Reading Speed Texts (IReST), which is a standardized paragraph reading task suitable for repeated measures and inter-language comparison of results (43). The IReST contains 10 parallel texts in newspaper print size (letter size 1M, or 10-point Times New Roman font). In the current study, text 1 was used for T1, text 7 for T2 and text 10 for T3 (all texts from the same category, indicating an average difference of less than 10 words per minute). The outcome measure is Words Per Minute (WPM), calculated as [(correctly read words/reading time in seconds)*60].

Additionally, as a measure for silent reading speed, we digitally presented texts 5 and 9 of the IReST at T1 and T2, respectively. Parts of texts 3 and 2 were selected as practice texts. The texts were presented in five segments of 3 lines each on a small tablet (10.1 inch), text size 1.7 mm. The participant was instructed to read the segment silently and subsequently press the button on the screen to move to the next segment. The presentation time was recorded for each segment, subsequently, the presentation times of the five segments were summed. The measure ‘WPM-silent’ was calculated as [(amount of words of text/sum of reading time of 5 segments in seconds)*60].

##### Self-reported Reading

The Hemianopia Reading Questionnaire (HRQ) was administered as a measure of self-reported relationship to reading, reading skills, daily life functional reading of different objects, reading time and reading goals for training. The HRQ is a novel questionnaire on reading specifically designed for adults with HVFDs (26). The HRQ inquires about current functioning and retrospective functioning regarding the time before the HVFD, which we labeled as time measurement ‘T0’. The HRQ was administered at T1, T2 and T3. The outcome measures are the subscale scores of the subscales Relationship to reading (5 items), Reading skills (8 items) and Reading objects (11 items). Our previous studies have uncovered indications that the HRQ subscale Relationship to reading consists of two separate subscales reflecting reading efficacy and reading attitude (Tol et al. (2024). Therefore, we will provide measures for these potential subscales instead, by calculating the mean score of items 1 and 4 (efficacy) and items 2, 3 and 5 (attitude).

##### Vision-related Quality of Life

We administered the 25-item National Eye Institute Visual Function Questionnaire (NEI-VFQ-25) as measure for vision- and health-related quality of life (44,45). This instrument is recommended for stroke-related visual impairment research (46). The NEI-VFQ-25 was administered at T1, T2 and T3, and the outcome measure was the composite score.

##### Control Measures

To control for any occurrences during or after the trainings or waiting period that would have impacted the outcomes in a major way, we included three control measures: 1) a custom visual field test at T2, 2) a post-training questionnaire at T2 and T3 and 3) training session reports filled in by the occupational therapists. The visual field test was a custom made 2-LT test that can be loaded onto both the Humphrey and Octopus visual field analyzers. The custom test consists of 68 visual stimuli within the central 10 degrees of the visual field, with 44 stimuli on the affected side, 10 stimuli on the midline and 14 stimuli on the unaffected side (see for a visual example Supplementary file S2). The custom test was administered at T2 to ensure no changed central visual field could be the explanation of the outcomes. The custom test at T2 was compared to the visual field perimetry administered in the diagnostic phase before T1, which was either done with ‘standard’ 10/2 central perimetry or with the custom test as well. The post-training questionnaire administered at T2 and T3 inquired about self-reported changes in vision, neurological status, major life events and whether another HVFD-training was followed. The session reports were filled in by the occupational therapists after each session, focusing on the covered themes or goals within the session, the length of the session, any deviations from the protocol, whether the participant had completed the homework and the participant’s level of concentration and fatigue.

#### Interviews on Training Outcomes and Application in Daily Life

Semi-structured interviews were conducted by phone at T2 with all participants who had followed either Vistra or Rotated Reading training. The interviews were aimed to inquire about the participants’ experiences with the reading training and what they had gained from the training. A topic guide was created to ensure coverage of the pre-defined topics of interest, while allowing for discussion of other, related concepts that came up during the interviews (see Supplementary file S3).

### Analysis of Results

#### Quantitative Measures of Reading and Quality of Life

##### Group Effects

To explore the potential of the outcome of the Vistra and Rotated Reading trainings, the effects of time, group and the time*group interaction on the IReST, HRQ and NEI-VFQ-25 were statistically analyzed. Analyses were repeated measures anova and post-hoc independent sample or dependent sample T-tests (IReST) or with non-parametric alternatives Friedman’s test and post-hoc Mann-Whitney U and Wilcoxon Signed Rank test (HRQ and NEI-VFQ-25). Due to the relatively small sample sizes, the estimation of the effectiveness of the Vistra and Rotated Reading trainings in the current study becomes more exploratory in nature. Therefore, we would rather accept the risk of a type 1 error than a type 2 error, and thus we did not apply a correction for multiple testing on the p-value. We did incorporate post-hoc power analysis and effect size calculation for the interpretation of effectiveness. Post-hoc power analysis was performed in the software G*Power (version 3.1.9.7) for the comparisons yielding a significant effect at *p* <.05. Sufficient power was regarded as a value of .8 or higher. Different effect sizes were calculated based on the corresponding statistical test; partial eta squared (*η^2^*) was calculated for repeated measures anova, Cohen’s *d* for T-tests, Kendall’s *W* for Friedman’s test, eta squared (*η^2^*) for Mann-Whitney U tests, and effect size *r* for the Wilcoxon Signed Rank test. The interpretation of the effect sizes was according to Cohen (1988); for the *η^2^* and *η^2^*, this is .01=small effect, .06=medium effect, .14=large effect. For Cohen’s *d,* Kendall’s *W* and effect size *r,* the interpretation is 0.2=small effect, 0.5=medium effect, 0.8=large effect.

##### Individual Changes

Individual change on the outcome measures word per minute, words per minute silent, reading efficacy, reading attitude, reading skills, reading objects and quality of life from T1 to T2 were assessed by calculating a Reliable Change Index (RCI). The RCI is a measure of individual statistical significance (48,49). The RCI was calculated with the following formula:

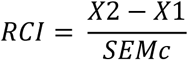

where X2 is the score on T2, X1 is the score on T1 and SEMc is the standard deviation of the difference score between T1 and T2 of the control group. This formula was chosen under the assumption that the control group reflects a condition of no true change. Furthermore, other variables to calculate the RCI with different formulas were not available (e.g. the reliability coefficient of the instruments) (48,49). As a RCI can be considered a z-value, a value of >1.96 was considered as a significant change. As we did not collect data in the control group at T3, a RCI for the difference between T2 and T3 was not calculated.

#### Interviews on Training Outcomes and Application in Daily Life

Transcripts of the interviews were created in the speech-to-text software Amberscript (2024). Transcriptions were analyzed by authors ST and JV by means of both inductive and deductive thematic analysis as described by (51). Analysis was performed in the qualitative data analysis software ATLAS.ti Web (version 9.2.0). Before coding started, deductive themes were formulated based on the research questions, such as ‘benefits of the training’ and ‘facilitating factors for implementation of the learned strategies’. Based on these initial themes and including any emerging inductive codes, both ST and JV independently coded two different interviews (one Vistra and one Rotated Reading). Based on these four interviews, an initial codebook was created in a discussion by ST and JV. Using this codebook, ST and JV each coded half of the interviews, while still allowing for any emerging inductive codes. After all interviews were coded, a final codebook was created. Using this final codebook, ST and JV recoded the interviews, each coding the interviews previously coded by the other. In this way, every interview was coded at least once by every coder. Afterwards, the codes from ST and JV were combined. Based on these combined codings of the transcripts, the codebook was analyzed in a discussion by ST and JV to create final codes and themes. Additionally, when the data was available, codes resulting from the interviews were verified against the quantitative items collected by providing frequencies of responses of the corresponding items on the HRQ.

Reflexivity statement: ST is a PhD student with background in clinical neuropsychology studying the potential effectiveness of reading training for individuals with HVFDs. She is particularly interested in the different range of potential outcomes of the trainings. Therefore these themes have been incorporated in the interview and initial codebook, as she extensively contributed to the design of the clinical trial and topic guide for the interview. Because of her background, particular attention might be steered towards psychosocial elements within the data. Additionally, ST has been involved in the development of the HRQ. The division of themes within the subscales of the HRQ may have potentially influenced her in analyzing the division of themes in the current study. JV is a master student studying Applied Cognitive Neuroscience and novel to compensatory reading training for individuals with HVFDs. She has an interest in HVFDs and previous experience with coding of interviews regarding reading with dyslexia. Because of her relative naivety to the topic, she is not expected to steer the analysis to a particular point.

## Results

### Quantitative Measures of Reading and Quality of Life

#### Group Comparisons

##### Words Per Minute

An overview of all statistical analyses can be found in Supplementary file S4. In Figure 2, the WPM for every group at every time point is shown. For participants in Vistra, a significant improvement of WPM with large effect size and sufficient power was observed from T1 to T2 (t(4)=−3.96, *p=*.017, *d*=−1.772, 1-β=.84). The control participants did not improve significantly on reading speed from T1 to T2. This is reflected in the significant interaction effect of time*group, indicating that the participants within the Vistra and control groups changed differently on reading speed before and after training (F(1,13)=4.86, *p*=.046, *η^2^* =.272, 1-β=1). Reading speed of participants in the Vistra group was also significantly lower compared to the waiting list control participants with large effects size on both T1 and T2 (t(13)=−4.05, *p*<.001, *d*=−2.216, 1-β=.96 and t(13)=−2.481, *p=*.028, *d*=−1.359, 1-β=.63). No significant effects for all comparisons were observed on WPM-silent. Furthermore, no effects on WPM were found for the Rotated Reading group.

**Figure 2.**
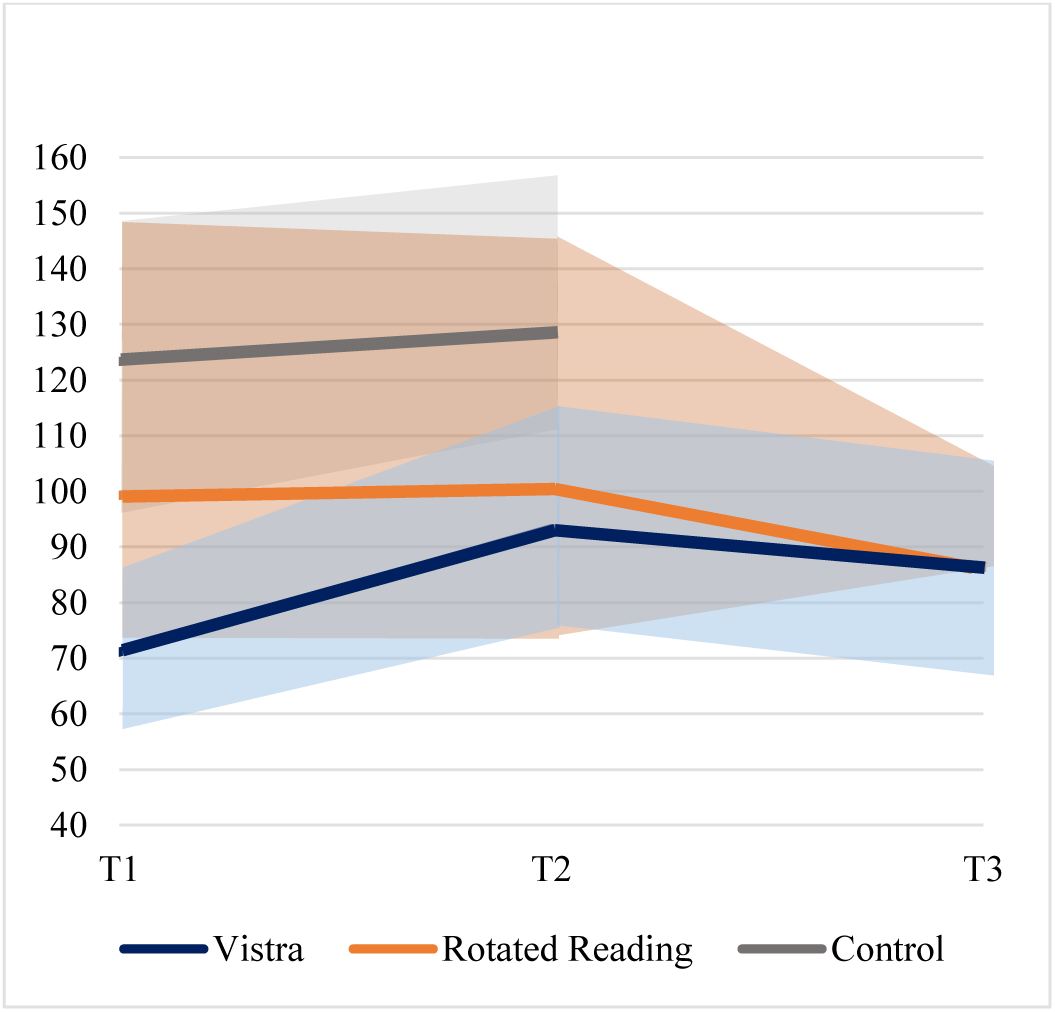
Group results on words per minute on the IReST reading task. *Note.* Error bars based on +/− standard deviation.

##### Self-reported Reading

The individual and group descriptive statistics of the HRQ subscale scores per group can be found in Supplementary file S5. An overview of all statistical comparisons made can be found in Supplementary file S4. At T1, participants were also asked to reflect on the items keeping the time before the HVFD in mind (‘T0’). For all groups, significant differences were observed between T0 and all other time points (T1, T2 and T3) on the HRQ subscales Reading skills and Reading objects. Thus, reported reading skills and reading of different objects never reached the perceived pre-HVFD level. On the items reflecting reading efficacy, significant differences compared to T0 were found on T1 and T3 in the Vistra and Rotated Reading groups and on T1 and T2 in the control group. Lastly, the scores on the items reflecting reading attitude were statistically similar for all groups on all time points.

No within-group differences were identified between T1 and T2 or between T2 and T3 on the subscales ‘efficacy’ and ‘attitude’ of HRQ Relationship to reading (see Supplementary file S4 and Figures 3a and 3b). Additionally, no differences between the training groups and control group were found at all time points on these subscales. Regarding self-reported reading skills (HRQ Reading skills), participants in both the Vistra and Rotated Reading groups showed a significant improvement with large effect size at both T2 and T3, compared to T1 (Vistra: T1-T2: Z=−2.03, *p*=.042, *r*=−.91, 1-β=.99; T1-T3: Z=−2.03, *p*=.042, *r*=−.91, 1-β=1; Rotated Reading: T1-T2: Z=−2.03, *p*=.042, *r*=−.91, 1-β=.73; T1-T3: Z=−2.02, *p*=.043, *r*=−.90, 1-β=.30; Supplementary file S4; Figure 3c). Note however that for the Rotated Reading group the power of these analyses is insufficient. A stable effect after training for both training groups, is indicated by no significant difference between T2 and T3. The control group did not report a significant difference in reading skills at T2, compared to T1. With regard to functional reading of different objects (HRQ Objects), the Vistra group showed significant improvement with large effect size on T2 and T3, compared to T1 (T1-T2: Z=−2.02, *p*=.043, *r*=−.90, 1-β=.98; T1-T3: Z=−2.02, *p*=.043, *r*=−.90, 1-β=1; Supplementary file S4, Figure 3d). Furthermore, the difference score between T1 and T2 differed significantly with large effect size between the Vistra and control group, indicating that the scores on T1 and T2 changed differently for the two groups (interaction effect; ΔT2-T1: U=2, *p*=.005, *η^2^*=.529, 1-β=.96). No such significant differences were found in the Rotated Reading group.

**Figure 3.**
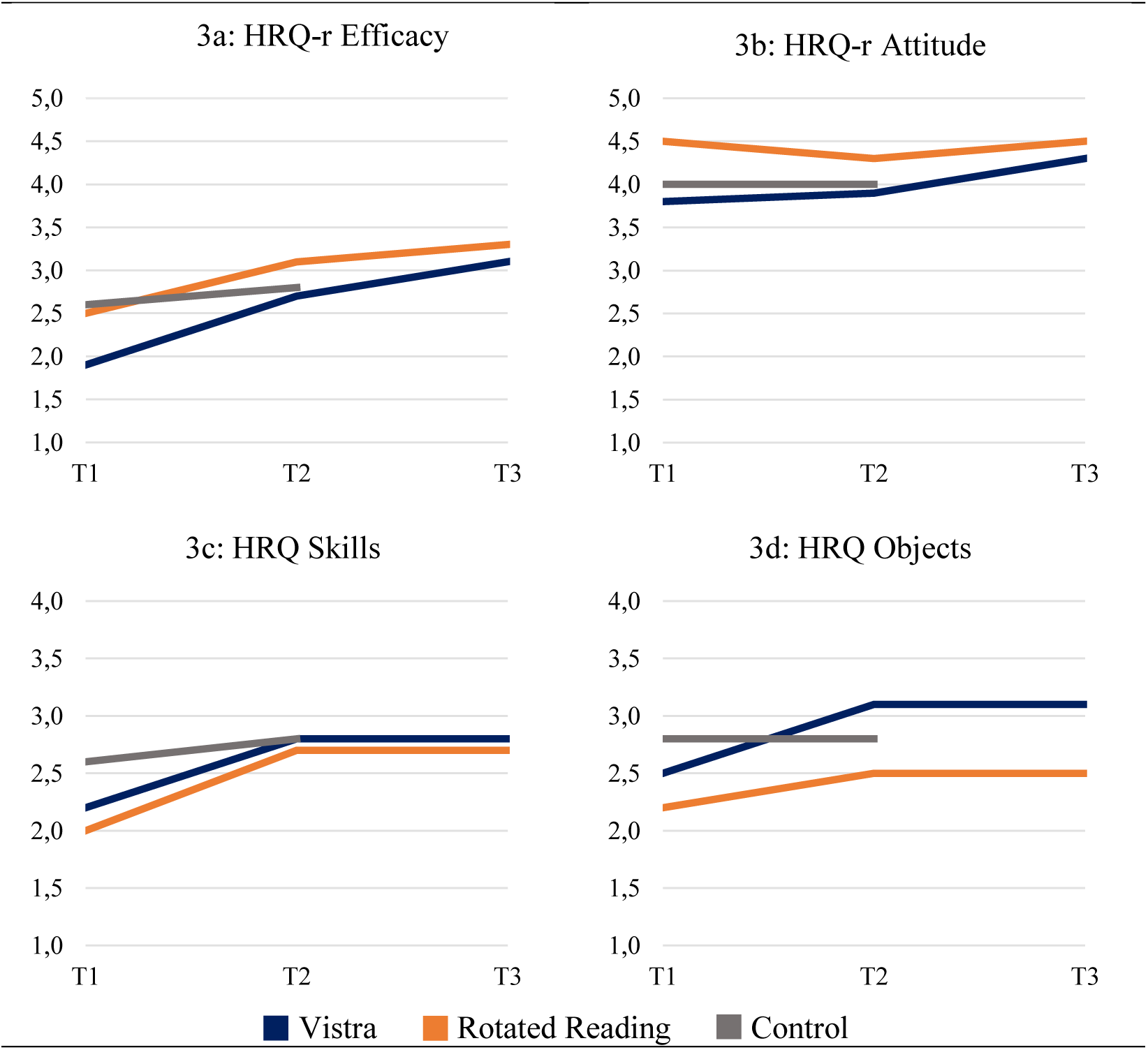
Group Results on the HRQ subscales Efficacy, Attitude, Skills and Objects at T1, T2 and T3. *Note.* Significant group effects were found for:

– HRQ Skills (Vistra T1-T2 and T1-T3, Rotated Reading T1-T2 and T1-T3, Vistra-Control at T1, Rotated Reading-Control at T1)
– HRQ Objects (Vistra T1-T2 and T1-T3, Vistra-Control interaction T1-T2, Rotated Reading-Control T1). See Supplementary file S4 for all statistical analyses, and Supplementary file S5 for the individual descriptive statistics of the HRQ data of the participants.

##### Vision-related Quality of Life

The interaction effect between the Rotated Reading group and control group appeared just significant with sufficient power (ΔT2-T1: U=9, *p*=.050, *η^2^*=.256, 1-β=.82), indicating that the scores between T1 and T2 changed differently for the groups. In Supplementary file S5 it can be observed that the vision-related quality of life in the Rotated Reading group decreased at T2, while the scores of the control group increased. No other statistical analyses with sufficient power indicated differences on vision-related quality of life (Supplementary file S4).

##### Control Measures

Due to changed visual fields at T2, two participants were excluded as mentioned in the Methods section. No other results on the post-training questionnaire and session reports were found that could have majorly impacted the outcomes. The completed session reports can be found in Supplementary file S6.

#### Individual Changes

The individual statistics of WPM of the participants from the training groups are presented in Table 3. The individual statistics of WPM-silent, HRQ subscales and NEI-VFQ-25 are presented in Supplementary file S5. Finally, the RCI for every outcome measure is presented in Table 4. Within the five participants from the Vistra group, one significant individual positive change was observed in reading speed read out loud, one in self-reported reading efficacy, three in reading objects and two in vision-related quality of life. Within the five participants from the Rotated Reading group, two significant individual positive changes were found on self-reported reading efficacy and one in reading skills.

**Table 3.** Descriptive Statistics of Words Per Minute.

| Participant | T1 | T2 | T3 | WPM |  |  |
| --- | --- | --- | --- | --- | --- | --- |
|  |  |  |  | *T2-T1* | *T3-T2* | *T3-T1* |
| V mean (N=5) | 71.11 | 93.29 | 86.63 | +22.18 | -6.66 | +15.53 |
| V1 | 62.69 | 102.35 | 86.25 | +39.66 | -16.1 | +23.56 |
| V2 | 58.71 | 80.75 | 91.01 | +22.04 | +10.26 | +32.30 |
| V3 | 60.00 | 64.48 | 57.43 | +4.48 | -7.05 | -2.57 |
| V4 | 88.42 | 108.75 | 85.53 | +20.33 | -23.22 | -2.89 |
| V5 | 85.71 | 110.13 | 112.94 | +24.42 | +2.81 | +27.23 |
| R mean (N=5) | 99.26 | 100.41 | 86.26 | +1.15 | -14.15 | -13.01 |
| R1 | 137.70 | 122.54 | 110.40 | -15.16 | -12.14 | -27.30 |
| R2 | 101.20 | 84.71 | 97.86 | -16.49 | +13.15 | -3.34 |
| R3 | 52.50 | 65.91 | 67.44 | +13.41 | +1.53 | +14.94 |
| R4 | 46.43 | 60.43 | 73.39 | +14.00 | +12.96 | +26.96 |
| R5 | 158.49 | 168.46 | 82.20 | +9.97 | -86.26 | -76.29 |
| C mean (N=10) | 123.35 | 128.23 |  | +4.88 |  |  |

*Note.* All participants had a macular sparing of less than 5 degrees.
WPM=Words Per Minute, HVFD=Homonymous Visual Field Defect,
V=Vistra, R=Rotated Reading, C=Control.

**Table 4.**
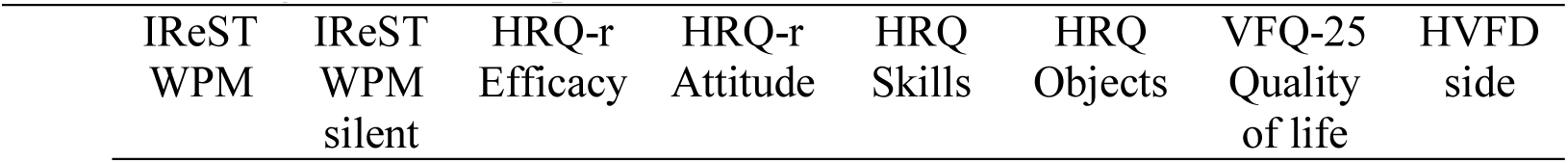
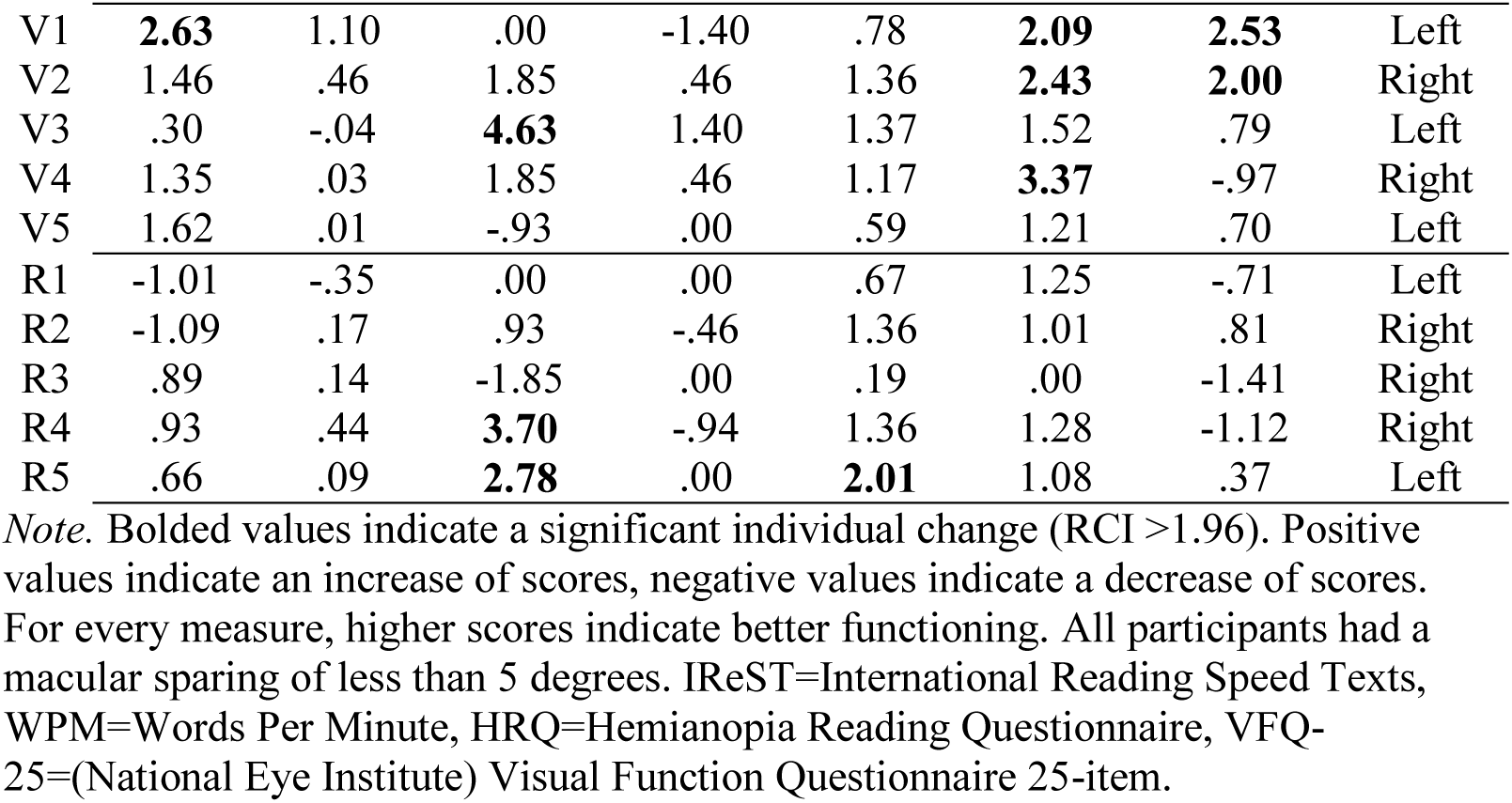
Reliable Change Index (RCI) per Outcome Measure for T2-T1.

| IReST<br>WPM | IReST<br>WPM<br>silent | HRQ-r<br>Efficacy | HRQ-r<br>Attitude | HRQ<br>Skills | HRQ<br>Objects | VFQ-25<br>Quality<br>of life | HVFD<br>side |
| --- | --- | --- | --- | --- | --- | --- | --- |

|  |  |  |  |  |  |  |  |  |
| --- | --- | --- | --- | --- | --- | --- | --- | --- |
| V1 | **2.63** | 1.10 | .00 | -1.40 | .78 | **2.09** | **2.53** | Left |
| V2 | 1.46 | .46 | 1.85 | .46 | 1.36 | **2.43** | **2.00** | Right |
| V3 | .30 | -.04 | **4.63** | 1.40 | 1.37 | 1.52 | .79 | Left |
| V4 | 1.35 | .03 | 1.85 | .46 | 1.17 | **3.37** | -.97 | Right |
| V5 | 1.62 | .01 | -.93 | .00 | .59 | 1.21 | .70 | Left |
| R1 | -1.01 | -.35 | .00 | .00 | .67 | 1.25 | -.71 | Left |
| R2 | -1.09 | .17 | .93 | -.46 | 1.36 | 1.01 | .81 | Right |
| R3 | .89 | .14 | -1.85 | .00 | .19 | .00 | -1.41 | Right |
| R4 | .93 | .44 | **3.70** | -.94 | 1.36 | 1.28 | -1.12 | Right |
| R5 | .66 | .09 | **2.78** | .00 | **2.01** | 1.08 | .37 | Left |

*Note.* Bolded values indicate a significant individual change ( $RCI > 1.96$ ). Positive values indicate an increase of scores, negative values indicate a decrease of scores. For every measure, higher scores indicate better functioning. All participants had a macular sparing of less than 5 degrees. IReST=International Reading Speed Texts, WPM=Words Per Minute, HRQ=Hemianopia Reading Questionnaire, VFQ-25=(National Eye Institute) Visual Function Questionnaire 25-item.

### Interviews on Training Outcomes and Application in Daily Life

#### Training Outcomes

The code tree for outcomes of the trainings as mentioned in the interviews can be found in Figure 4. Additional to identifying codes and themes regarding training outcome, we could distinguish whether the participants experienced positive improvement, a bit improvement or no improvement on the mentioned outcomes. No participant mentioned any item that had deteriorated after reading training. In total, five overarching themes and three subthemes regarding training outcomes could be developed from the data, covering 25 individual codes. It is important to mention that Figure 4 represents potential outcomes for the iwHs who have followed reading training. The qualitative data does not provide an indication on the frequency of the outcomes for the full population of iwHs. We have added the individual data of the HRQ items in the following section in order to compare the subjective spontaneous (interviews) and standardized (HRQ) outcomes.

**Figure 4.**
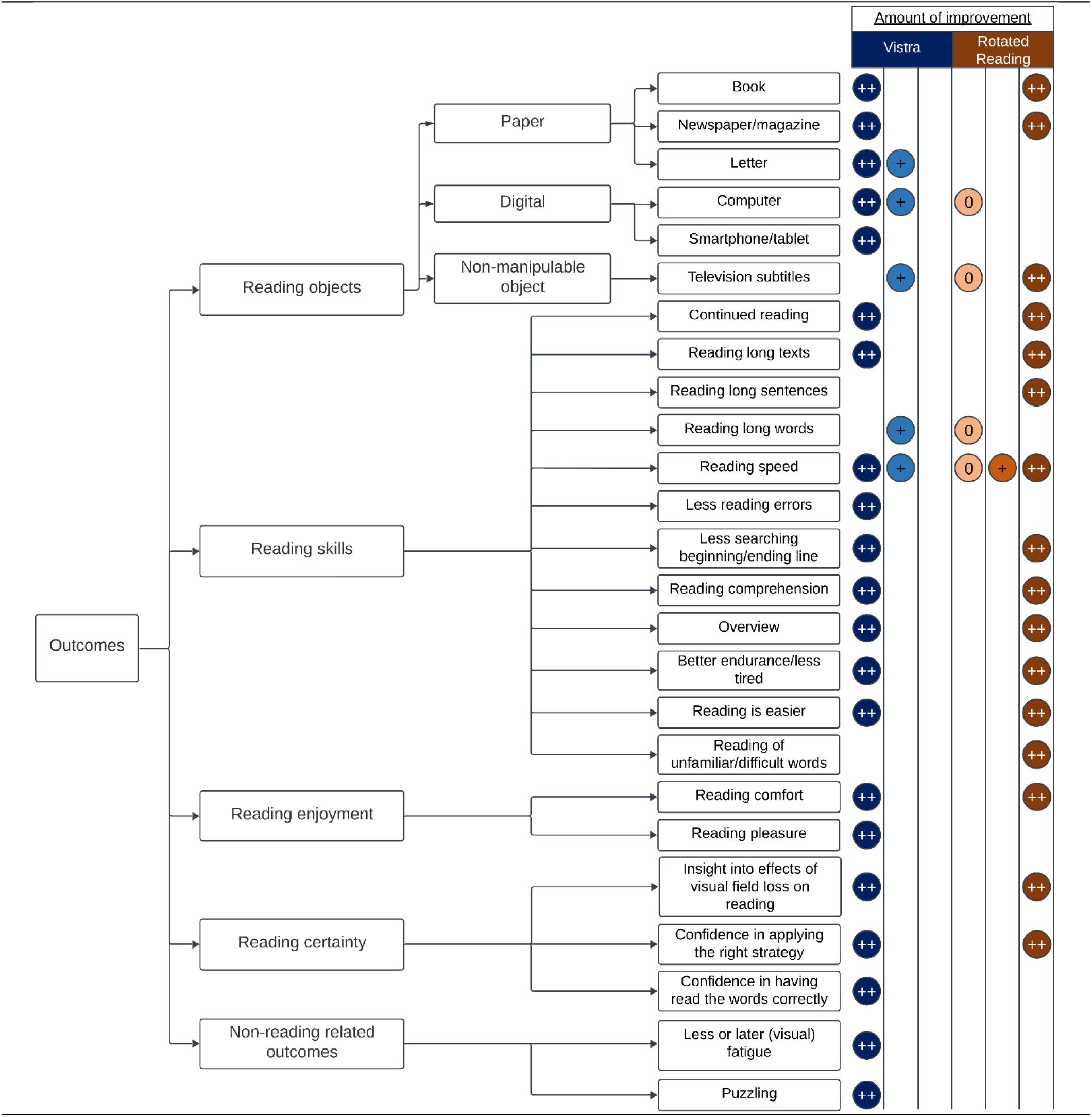
Code Tree of Spontaneously Mentioned Training Outcomes. *Note.* ‘++’ indicates a positive improvement, ‘+’ indicates a bit positive improvement, ‘0’ indicates no improvement.

##### Theme 1: Reading Objects

This theme is divided into three subthemes; paper objects, digital objects and non-manipulable objects that are far away (i.e. television subtitles). The theme describes potential improvement of reading of six different objects that participants have mentioned in the interviews. Therefore, in Table 5 the change in responses on items on the HRQ Objects subscale is provided, indicating whether participants reported a different score (lower, same or higher). It can be observed that for the Vistra training, improvement at T2 was reported most often (4 out of 5 participants) on reading subtitles and from a smartphone. In the Rotated Reading training, improvement at reading a physical book was reported most often (4 out of 4 participants).

**Table 5.**
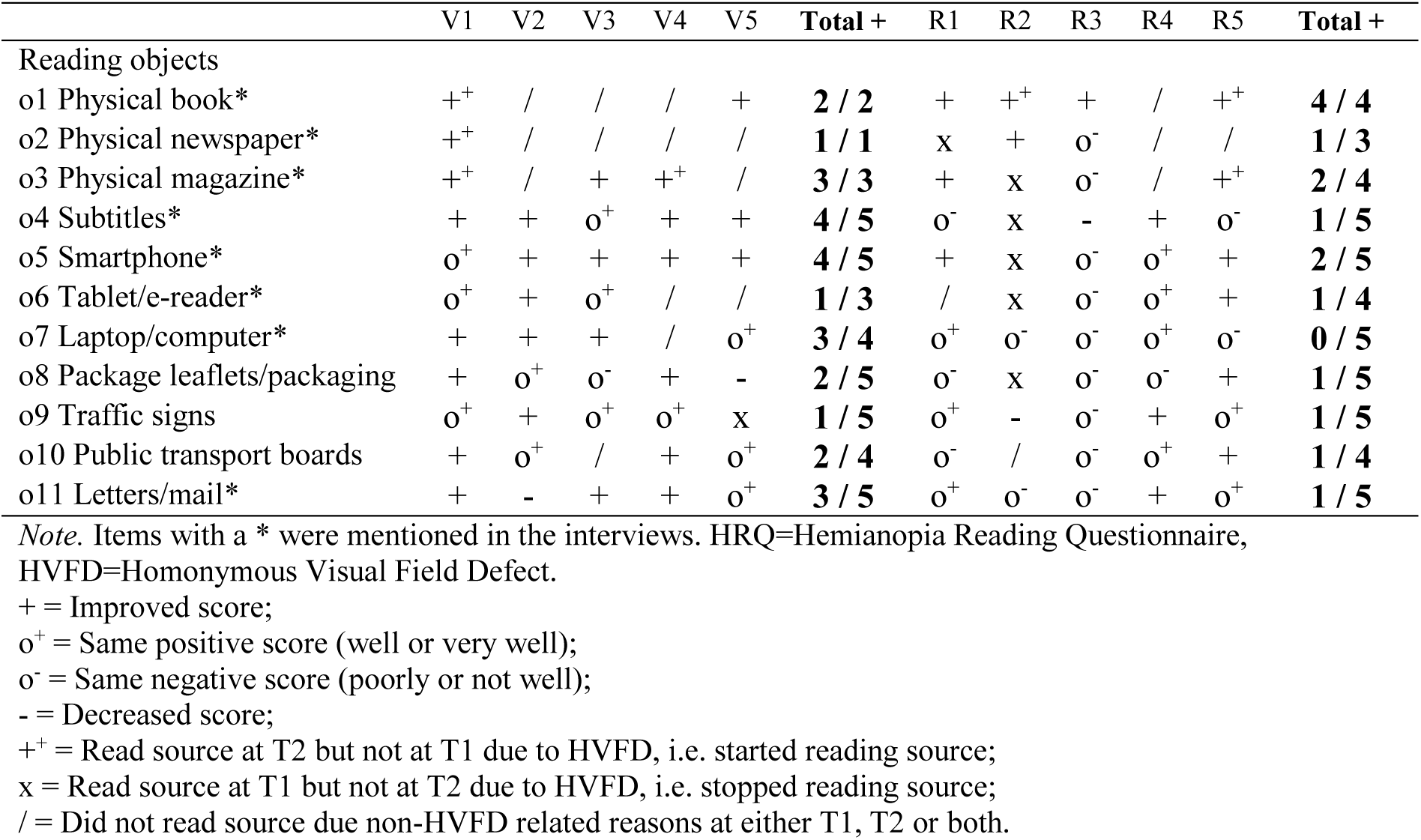
Change in Scores on the Items of the HRQ Objects Subscale From T1 to T2.

##### Theme 2: Reading Skills

The participants mentioned in total twelve reading skills that improved after reading training. Though all skills were mentioned as having been positively improved, some variation can be found in e.g. reading speed, where for both trainings ‘a bit improvement’ was also mentioned. For the Rotated Reading training, ‘no improvement’ was also mentioned in relation to reading speed. Often, the interplay between multiple skills improving was mentioned by participants (i.e. skills influencing each other). For example, one participant reflected on text rotation resulting in the improvement of multiple reading skills:

> *When I start reading an A4 page in the normal way, especially if there’s a lot of text on it, it takes me more time, I make more mistakes, I have to go back and look things up more, and then it actually just costs me more energy. [..] So when I’ve read, say, one or two A4 pages, then that’s it for me. And this way, so the rotated reading, it turned out that I get the entire piece of text into my view again. [..] making it easier, making fewer mistakes, having to look back less, costing less energy for me, and in the end I can take in the information better too. (Rotated Reading participant)*

Another rather interesting quote was provided by a participant from the Vistra group. It concerns a description of improved reading endurance, which seems to present as no longer experiencing the onset of a tunnel vision during reading:

> *Before when I was reading then everything outside the bit I was reading went completely black at one point. That’s not the case anymore. Yes I think that was due to fatigue. That [tunnel vision] eventually emerged when I was reading and it was really like I was in a dark tunnel with a little bit of light on which I was focusing in the middle [..]. Researcher: And you don’t experience that anymore? Or less? Participant: No, not anymore actually. That could be because now I’m able to read maybe an hour at a time so to speak. I haven’t tried it out, but I could do it. Maybe it [tunnel vision] will happen after an hour, where before it was after four minutes.. (Vistra participant)*

Table 6 shows the individual changes in responses from T1 to T2 on items from the ‘efficacy’ and ‘reading skill’ subscales of the HRQ. Here it can be observed that the participants experienced an improvement in most reading skills. All participants in the Vistra group, responded with a higher score on T2 compared to T1 regarding being able to locate the next line. Furthermore, 4 of out 5 participants from this group indicated a higher score regarding, perceiving a long word, reading without fatigue, remembering what is read stand and experiencing no difficulty with reading. For the Rotated Reading training, understanding what is read, fast reading and experiencing no difficulty with reading.were the reading skills in which improvement was mentioned the most (in 4 out of 5 participants) are

**Table 6.** Change in Scores on the Items of the HRQ-r Efficacy and HRQ Skills Subscales Regarding Reading Skills From T1 to T2.

|  | V1 | V2 | V3 | V4 | V5 | Total + | R1 | R2 | R3 | R4 | R5 | Total + |
| --- | --- | --- | --- | --- | --- | --- | --- | --- | --- | --- | --- | --- |
| Reading skills |  |  |  |  |  |  |  |  |  |  |  |  |
| s1 Understanding | o<sup>-</sup> | o<sup>+</sup> | + | - | - | **1 / 5** | + | + | + | o<sup>+</sup> | + | **4 / 5** |
| s2 Fast reading | o<sup>-</sup> | + | o<sup>-</sup> | + | + | **3 / 5** | - | + | + | + | + | **4 / 5** |
| s3 Locating next line | + | + | + | + | + | **5 / 5** | + | + | - | o<sup>+</sup> | + | **3 / 5** |
| s4 Finishing a line | o<sup>+</sup> | o<sup>+</sup> | + | + | + | **3 / 5** | + | o<sup>-</sup> | o<sup>-</sup> | + | + | **3 / 5** |
| s5 Perceiving a short word | o<sup>+</sup> | + | + | o<sup>+</sup> | - | **2 / 5** | o<sup>-</sup> | + | o<sup>+</sup> | + | + | **3 / 5** |
| s6 Perceiving a long word | - | + | + | + | + | **4 / 5** |  | + | o<sup>-</sup> | + |  | **2 / 3** |
| s7 Reading without fatigue | + | + | + | o<sup>+</sup> | + | **4 / 5** | o<sup>-</sup> | + | o<sup>-</sup> | + | + | **3 / 5** |
| s8 Remembering | + | o<sup>+</sup> | + | + | + | **4 / 5** | + | + | o<sup>+</sup> | - | + | **3 / 5** |
| Reading efficacy |  |  |  |  |  |  |  |  |  |  |  |  |
| r1 I am a good reader | o<sup>-</sup> | + | + | + | - | **3 / 5** | o<sup>+</sup> | o | - | + | + | **2 / 5** |
| r4 I experience no difficulty | o<sup>-</sup> | + | + | + | + | **4 / 5** | o<sup>-</sup> | + | + | + | + | **4 / 5** |

*Note.* Items with a \* were mentioned in the interviews. For participants R1 and R5, no was available on item 6. HRQ=Hemianopia Reading Questionnaire. HRQ-r=HRQ Relationship to reading subscale.
‘+’ = Improved score;
‘o<sup>+</sup>’ = Same positive score (HRQ-r Efficacy or strongly agree, HRQ Skills well or very well);
‘o’ = Same score (HRQ-r Efficacy neither agree or disagree or same amount of minutes);
‘o<sup>-</sup>’ = Same negative score (HRQ-r Efficacy strongly disagree or disagree, HRQ Skills poorly or not well);
‘-’ = Decreased score.

##### Theme 3: Reading Enjoyment

Participants mentioned reading comfort and reading pleasure having been positively improved after reading training. For example, one participant from Vistra group mentioned that the motivation for reading has increased after training:

> *I have a book here. Well, it’s been here since my birthday, since [6 weeks ago]. I once read one page in it, because after half an hour I thought, well I’ll stop, I’m exhausted you know. I have to go back every time [..] But that goes more smoothly now, so I notice that now I experience some yes, somewhat less of a barrier to pick up a book, to read. (Vistra participant)*

As can be seen in Supplementary file S5, a few individuals from both training groups showed increased scores at T2 on items regarding the importance of reading, a positive attitude towards reading and love for reading. However, most individuals had reported a positive score for these items already at T1 and these scores did not change at T2.

##### Theme 4: Reading Certainty

Overall, this theme describes certainty in reading that the participants have gained after reading training. The theme entails two codes referring to confidence that participants have gained after reading training in 1) applying the right strategy (mentioned for both reading trainings) and 2) in having read the words correctly (mentioned for Vistra). Additionally, this theme includes a code referring to insights into the effects of the visual field loss on reading. For example, two participants mentioned the following:

> *Otherwise you’re just trying something yourself, so then you don’t actually have an assurance of ‘I’m doing it the right way’. And that idea alone has helped me a lot. (Rotated Reading participant)*

> *That what I’m reading now, that that’s what it says, let’s put it that way. And I’m also reading the whole word now, I think or I’m pretty sure, because it can’t be that if I start at the beginning and I go to the end like this letter by letter, so to speak, I can’t go wrong. (Vistra participant)*

##### Theme 5: Non-reading Related Outcomes

Some participants also mentioned non-reading related outcomes of reading training. One code that could be created concerns an experienced improvement in puzzling (i.e. jigsaw puzzles) after training. The other code covers participants describing less or later overstimulation or fatigue during the day. An example was provided by a participant from the Vistra group:

> *Well I suspect that, I didn’t measure it, but before I was in a constant state of being overstimulated. Because of the [acquired brain injury]. And I feel that that has become less. [..], Look, if I get up in the morning, it depends a bit on the day before, but then I am not overstimulated. But during the day, the overstimulation starts to emerge. There is nothing you can do about it, it just happens. [..] I notice that I am less overstimulated, so not the whole day, but rather at the end of the day. (Vistra participant)*

#### Application in Daily Life

The code tree regarding learned strategies and their application after reading training can be found in Figure 5. This code tree represents learned strategies that the participants have mentioned spontaneously, as well as what would be needed to continue with these strategies (facilitating factors).

**Figure 5.**
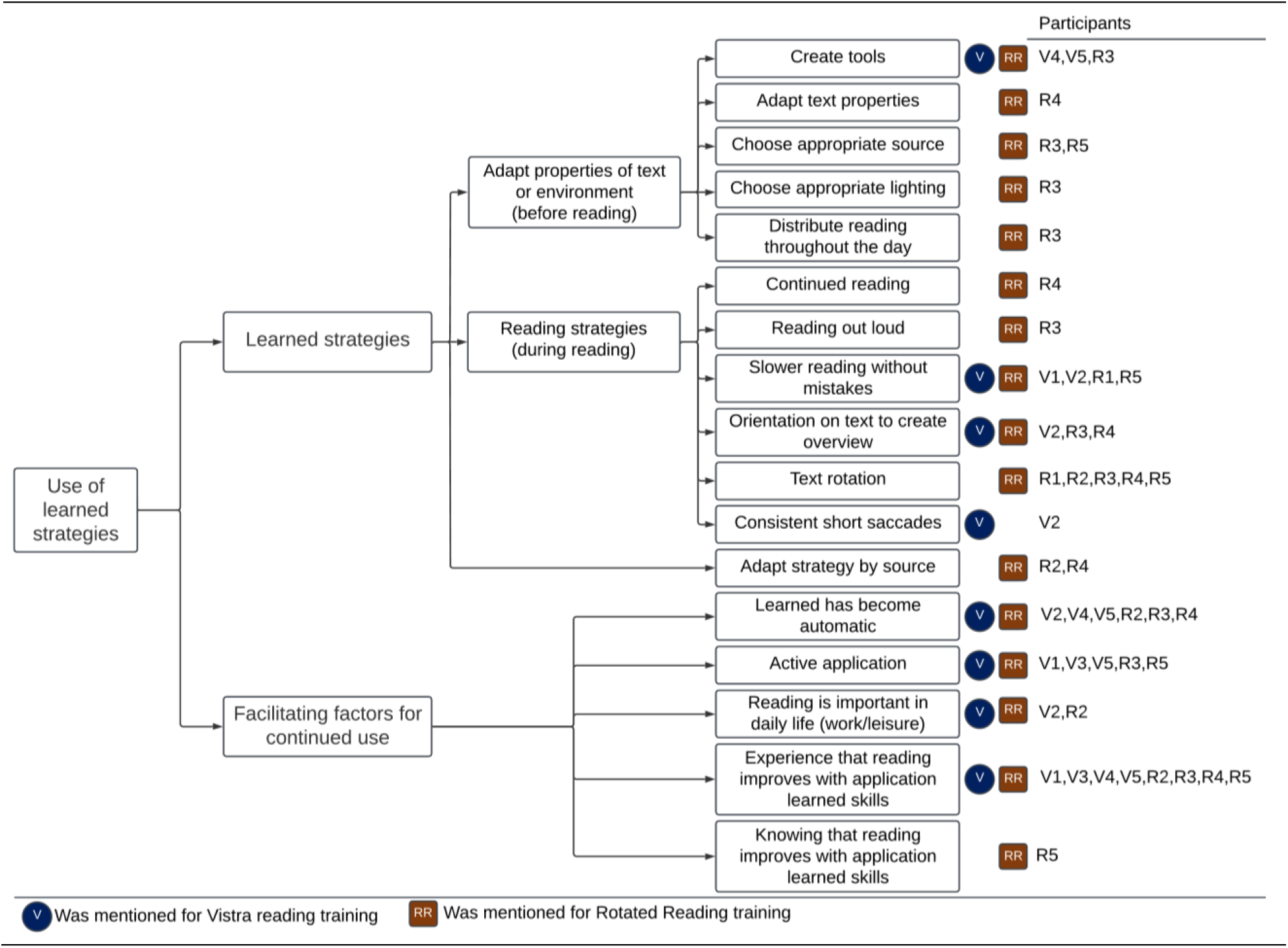
Code Tree of Spontaneously Mentioned (Use of) Learned Strategies.

##### Theme 1: Learned Strategies

The participants described strategies that they had learned during reading training, which could be divided into three categories. Firstly, the subtheme adapting the properties of the text or environment describes strategies that can be applied by the iwH in preparation for reading. For example, one participant described the use of a rotation tool:

> *And especially that it’s much easier because I have a paper that has a line on it and you have to put that [reading object] exactly against that line every time. That initiates it [rotated reading] as it were. (Rotated Reading participant)*

Participants also described choosing appropriate sources:

> *I notice this because I, yes, I do find it a bit clearer when I pick up a book now, then I take a really good look first: how is it, how is it printed, how are the distances? And then I start reading with more pleasure. (Rotated Reading participant)*

> *What has improved in that sense is that if I get a longer email, for example, and I print it out and I read it in the rotated way, that does bring improvement. (Rotated Reading participant)*

The second subtheme is applying reading strategies during reading. Most mentioned strategies were slower reading without mistakes, creating overview and text rotation. Lastly, participants mentioned adapting the learned skills dependent on the different reading objects, thus applying different strategies for different reading objects.

##### Theme 2: Facilitating Factors for Continued Use

This second theme includes codes mostly emerging from answers to the question: “What is needed to continue using what you have learned?” (see topic guide in Supplementary file S3). Codes relating to this theme were mentioned by participants from both training groups and largely overlapped. For both trainings it was mentioned that the learned strategies had already become automatic, while also within both training groups it was mentioned that active application of the learned strategies was still needed to continue using the strategies. Further, it was mentioned that reading being important for either work or leisure activities were facilitating factors to continue the learned strategies. Finally, either the experience or knowledge that reading improves with the learned strategies were mentioned as facilitating factors for continued use:

> *If you have a method, which is helpful, which is better than what you’ve been doing primarily, yes, that then, then you don’t drop it so quickly of course. (Rotated Reading participant)*

#### Non-applicability of Rotated Reading

Multiple situations in which specifically reading of rotated texts cannot be applied emerged from the interviews with the five participants who followed the Rotated Reading training (no such examples were mentioned by the Vistra group). These situations can be found in Table 7, and emerged mainly from participants reacting to the question: “What is a reason for you not to apply what you have learned?” (see topic guide in Supplementary file S3). A reported example of text rotation not being necessary when reading from an e-reader with adaptable screen properties is as follows:

> *Well, when I have a text in front of me with four words in a row and good line spacing on an uncluttered, small screen, well, that actually eliminates the problems I experience when reading a normal book. (Rotated Reading participant)*

**Table 7.** Situations When Rotated Reading is Not Applied.

|  | Mentioned by |
| --- | --- |
| Non-rotatable object (e.g. public transport/traffic signs or television) | R2, R3, R4 |
| Not necessary because text formatting can be adapted (e.g. e-reader) | R1, R4, R5 |
| Unable to gain easy overview of rotated text (e.g. pop-ups on computer or figures in newspaper) | R4 |

#### Feedback on Trainings

Information with regard to spontaneously mentioned feedback for the reading trainings was extracted from the interviews. The feedback is presented in Table 8. Multiple participants have expressed a desire for a personalized approach within reading training, by e.g. incorporating their own training materials or matching the training intensity and scope of the homework to the capacities of the participant. It should be mentioned that these elements are already part of both the Vistra and Rotated Reading protocols. Additionally, one participant has expressed to intend not to read digitally anymore since the HVFD, and therefore prefer to train with paper exercises only.

**Table 8.** Spontaneously Mentioned Feedback on Reading Training for Homonymous Visual Field Defects.

|  | Mentioned by |
| --- | --- |
| Flexibility with regard to paper or digital training | R5 |
| Follow-up session/aftercare | R5 |
| Share measurement of results of training with the participant | V1, R3 |
| Personalized approach: | V2, V3, V5, R1, R3, R5 |
| • With regard to the pacing of the training<br>• With regard to the materials:<br>- Materials of participant's own choice<br>- Materials that are sensitive to the native language of the participant<br>- Homework matched with participant on difficulty level |  |
| Include television subtitles in training | V2 |

## Discussion

In the current study, we assessed two compensatory reading trainings for iwHs that have been developed in Dutch clinical practice: Vistra and Rotated Reading. The aim was to explore the potential outcomes of the trainings and gather information for further optimization of reading training research and practice, by examining the outcomes and patient experiences. These insights inform two elements of EBP, namely scientific evidence on the effectiveness of training and patient perspectives (22). We found significant positive effects after Vistra on reading speed, self-reported reading skills and functional reading of different reading objects. No significant group effects with sufficient power of Rotated Reading training were found so far, but there are underpowered indications for an improvement in self-reported reading skills. Furthermore, for some individuals who had followed Vistra, significant individual changes were observed on reading speed, self-reported reading efficacy, daily life reading of different objects and vision-related quality of life. For individuals who had followed the Rotated Reading training, some significant individual changes were found on self-reported reading efficacy and reading skills. The current study offers a qualitative analysis on training outcomes resulting in five themes: reading objects, reading skills, reading enjoyment, reading certainty and non-reading related outcomes. Experienced learned skills in the trainings can be grouped into strategies in preparation to reading, strategies during reading and source-specific adaptations. The most mentioned motivating factors for continued use of the learned skills was the experience that reading has improved. For the Rotated Reading intervention, situations where text rotation is not applied were identified, suggesting limited suitability in distinct cases. Finally, feedback points for reading training highlighted predominantly the wish for personalized options within the trainings.

### Reading Speed

The most statistically robust group effect in our study was found on reading speed (WPM read out loud) after Vistra. A significant interaction indicates improvement in the Vistra and not in the control group. Reading speed did not significantly change between training and follow-up, indicating a stable effect. This corresponds to earlier saccadic reading training studies that all studied single-word reading tasks and showed a significant increase in reading speed after training (13,17–19). Other authors studied a saccadic compensation training for iwHs that combined three techniques (saccadic training, reading speed training and line reading training) (25). In this study an increase was found in text reading from both paper and a computer screen after training. Though these mentioned trainings and Vistra are not identical, a considerable overlap is evident in the training of reading eye movements. Thus, we conclude that, in line with previous studies on saccadic compensation trainings, Vistra seems a promising method to improve reading speed in iwHs. Furthermore, one of the five participants from the Vistra group showed a significant individual change after training.

We did not find significant effects of Rotated Reading training on reading speed. Some earlier studies have investigated the effect of rotated reading on reading performance in iwHs (11,20,21). Two of these studies have done so in the form of a home-based daily text rotation training, consisting of reading single lines (20,21). This training had a higher intensity compared to our Rotated Reading training, which was supervised at the rehabilitation centre in 5 or 6 weekly sessions (depending on the participant, see session reports in Supplementary file S6). Additionally, in the Rotated Reading training the rotation angle was individually determined based on the visual field and preference of the participant. Recent studies found that vertical reading training improved reading speed on the IReST post-training and at follow-up significantly (20,21). However, this effect was only significant in the subgroup analysis with individuals with right-sided HVFDs. In both these studies, a change of 10 WPM was deemed clinically relevant (which three of their 11 participants obtained). Of the five participants who followed the Rotated Reading training in the current study, three showed an increase in WPM post-training (+13, +14 and +10), while two showed a decrease (−15 and −17), though all not statistically significant according to the RCI measure. However, adhering to the criteria of Kuester-Gruber, all these changes would be considered clinically relevant. Because our small sample does not allow further subgroup analysis, it is difficult to examine why two participants showed a decrease in reading speed post-training. These two participants showed rather high reading speeds before training (101 and 138 WPM). Potentially, these participants have received the advice in training to read more slowly, but accurately, as the code ‘slower reading without mistakes’ could be created from the interviews regarding learned strategies. Furthermore, there seems to be a considerable difference between the WPM before training between the Vistra (71 WPM) and Rotated Reading (99 WPM) groups, potentially leaving less room for improvement in the Rotated Reading group. However, earlier mentioned studies that found positive significant effects after reading training did include groups with pre-training reading speeds of 90-96 WPM (13,17,19,53). Future research is needed to further uncover the benefits of text rotation training for iwHs on reading speed.

Unsurprisingly, silent reading speed was considerably higher for all participants, compared to reading speed when reading out loud (54). No significant effects of training on silent reading speed were found. It is interesting to note the discrepancy between read out loud reading, on which we found some group and individual effects after training, and silent reading, where we found no significant effects. Silent reading will be typically the most common form of reading in daily life. However, it will probably be affected in the same way as reading out loud is by HVFDs. It must be noted that we have no information about the validity of the digital segmented IReST as assessed in the current study.

### Self-reported Reading and Vision-related Quality of Life

Based on our small sample, Vistra seems a promising method to increase self-reported reading skills and reading of different reading objects. Notably, three out of five participants even showed a significant individual increase on reading of different objects. Earlier researchers have argued that, as statistically significant group results may occur without any individual significant changes within that group, the clinical relevance of treatment outcomes might be better reflected in the amount of individuals with significant change within that group (48). Thus, our findings might indicate a clinically relevant positive effect of Vistra on reading of different objects. Additionally, one participant showed a significant individual change in self-reported reading efficacy. For the Rotated Reading training we found indications (i.e. significant with large effect size, but underpowered) for improved self-reported reading skills in iwHs. Regarding the Reading attitude subscale of the HRQ, a positive and stable response in all three groups on all measurements (including the time before the HVFD - retrospectively reported), was found. Thus, both the occurrence of HVFD and reading training do not influence reading attitude. This finding builds upon our earlier finding that iwHs show no different scores on these reading items of the HRQ compared to matched individuals without HVFDs (55). These findings indicate the importance and love of reading are consistent, and thus emphasizing the relevance of reading trainings for iwHs.

Compared to other samples from HVFD-intervention studies, our sample showed similar pre-training scores with regard to quality of life as measured on the 25-item or 39-item NEI-VFQ (56–58). Unlike findings in earlier studies, we did not find any group-based indications for an improvement in vision-related quality of life after both reading trainings. However, we did find individual significant increases in vision-related quality of life at T2 in two participants from the Vistra group.

### Notable Individual Observations: Reading and Cognitive Fatigue

Most of our participants indicated being able to read longer without fatigue after reading training, which indicates a better experienced reading endurance. Furthermore, two participants experienced less overstimulation after Vistra, expressed in a delayed feeling of overstimulation during the day, and a famished narrowing tunnel vision during reading, respectively. Reading is a complex cognitive activity (59), and especially cognitively demanding in individuals with brain injury (60). We hypothesize that improved reading skills after reading training may result in less overall cognitive fatigue, which in turn postpones other expressions of cognitive fatigue such as the feeling of being overstimulated or experiencing visual complaints like an experienced tunnel vision. Some earlier studies have suggested a relationship between cognitive fatigue and visual complaints in individuals with acquired brain injury (61–63). Following such observations, other authors have shown in their study the potential of reading training for reducing fatigue during reading (25). Our results further support this idea. Furthermore, we hypothesize that, for some individuals, the benefits of reading training might even surpass reading-related fatigue and generalize towards other expressions of cognitive fatigue as were mentioned in the interviews. However, to the best of our knowledge, such hypotheses have not been studied.

### Clinical and Scientific Implications

Taking together all sources of information, positive outcomes were found for both trainings in which iwHs learn to adapt their reading behavior in 5 (Rotated Reading) or 10 (Vistra) sessions. Even with a small sample size, statistical group effects were found of Vistra on reading speed, self-reported reading skills and reading objects. Comparable to our study, other authors performed a small sample study on HVFD-trainings and included quantitative measures as well as interviews (64). They found limited statistical effects on the quantitative measures but did find, as did we, a range of experienced positive outcomes based on interviews after intervention. Therefore, these authors concluded that scarcity of quantitative significant effects can not be interpreted as an indication of no effect, as the qualitative outcomes contradict this. We align our perspective on the current outcomes with this observation by Hazelton and colleagues. In Table 9, we present some specific reading goals for which we recommend the Vistra or Rotated Reading training, based on the current results.

**Table 9.**
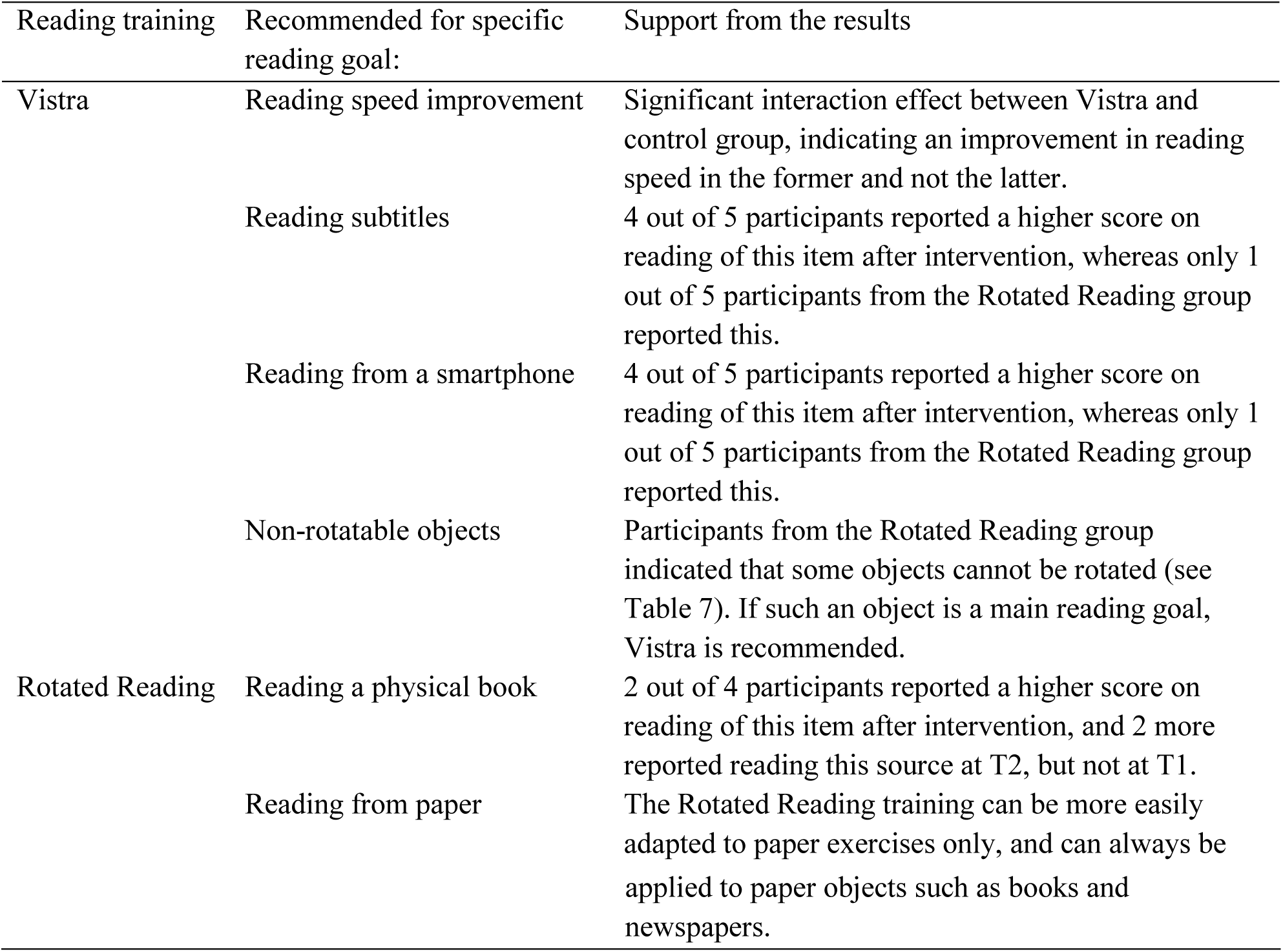
Initial Recommendations for Vistra and Rotated Reading.

Multiple participants mentioned the wish for personalized options in the reading trainings regarding for example the training pace or difficulty of homework assignments. Additionally, bringing one’s personal reading materials to the training was mentioned multiple times, which might offer some advantages. Personal reading materials match most closely the objects that the iwH will read in daily life after the reading training, might therefore invoke the most interest during the training, and offer the opportunity to incorporate texts in the native language of the participant. Some reading training studies for iwHs have already incorporated the use of personal texts (20,21,65). Another observation from the interviews concerns a wish for training with paper materials, due to self-reported screen sensitivity/intolerance after brain injury. There is, however, a growing body of literature on telerehabilitaion, which offers multiple benefits as access to rehabilitation for individuals who are not able to attend regular care due to physical, economical or geographical constraints (66,67). Also in HVFD reading training research, some authors have focused on telerehabilitation (20,21,64,65,68,69). Nevertheless, it is good to keep in mind the potential subgroup of iwHs who prefer to (or only can) read from paper. Examples of paper-based saccadic reading trainings for iwHs are the Brainwave-R workbook and Rainbow Readers (64,70).

### Limitations

A few limitations of the current study should be mentioned. First, we did not register whether and in which angle participants from the Rotated Reading group rotated the IReST at T2 and T3. The participants were instructed to rotate the paper if they had learned this in the training. An earlier study has shown that reading speed improvement seems to be exclusive for the direction in which was trained (horizontal or vertical) (21). Therefore, to better understand the effects of the Rotated Reading training on reading speed, it would have been informative to register if the participants did actually rotate the paper, and if this was in the same angle as they had practiced within the training. Second, due to privacy regulations, we are not able to provide individual information about the participants other than the side of the HVFD. Due to their suggested effect on reading and HVFD-intervention in iwHs, it would have been interesting to also include individual information about the size of the HVFD, the amount of macular sparing and time since lesion (2,9,14,42). Last, including more participants would have allowed for more representative and generalizable comparisons with higher statistical power. Additionally, as the RCI measure was calculated with the scores of the control participants as norm reference, a larger sample of control participants would have been desirable to enlarge the representativeness of the control group scores as the norm in the RCI calculation. Furthermore, a larger sample would have allowed for sub-group analyses, which would have been informative to understand which training might suit which iwH the best. Sub-group analysis could again be focused on HVFD characteristics such as side and size of the HVFD and the amount of macular sparing. Earlier studies on saccadic compensation reading training found no different outcomes for individuals with left-sided or right-sided HVFDs (17,19), though two studies did find that individuals with right-sided HVFD needed more trainings sessions (18,71). However, there are some indications that vertical text reading is primarily beneficial for individuals with right-sided HVFDs (20,21). Furthermore, it could be hypothesized that the Rotated Reading training might be especially suitable for individuals with quadrantanopia or partial central field loss, as a minimal angle of rotation is required (just a few degrees into the particular sparing of the central visual field). This results in a smaller alteration of the visual representation of the word one has to read. However, based on our relatively small sample, no clear indications for an effect of HVFD side, size or macular sparing on the reading outcome measures could be found for both trainings.

## Conclusion

The current study set out to explore the potential effectiveness of the reading trainings Vistra (saccadic compensation) and Rotated Reading (text rotation), which have been developed in Dutch clinical practice for iwHs. The use of a mixed-methods approach including objective and subjective measures of reading as well as qualitative methods has allowed us to create a comprehensive notion of the potential effect of the reading trainings Vistra and Rotated Reading. The different items of the HRQ allow for a detailed overview of the changes after training in reading attitude, efficacy, skills and reading of different objects. Furthermore, these results provide insight into where the potential benefits of the reading trainings lie for each individual studied. This can be relevant, as significant group effects are no guarantee for the effectiveness of any training for the individual (22,48). The current results indicate a group effect of Vistra on increased reading speed (read out loud), self-reported reading skills and reading of different objects. For both Vistra and Rotated Reading, a range of individual positive outcomes were found based on individual significant changes on the tests and on the interviews. Notably, more than half of participants expressed the desire to personally tailor reading training. More research is Our warranted with larger study samples to ensure generalizable data and to allow for sub-group analyses. Based on the current results, we advise to incorporate the reading goals regarding the situations in which one wants to read and objects one wants to improve on when selecting Vistra or Rotated Reading training.

## Data Availability

Data cannot be shared publicly because of privacy sensitivity of the data. Anonimized data can be shared upon reasonable request by Gera de Haan from the Univserity of Groningen.

## Acknowledgements

This research was funded by ZonMw, project no. 94313005 and and ZonMw, programme Expertisefunctie Zintuiglijk Gehandicapten, grant number 637005001. First, we would like to thank our participants as without them, no clinical research is possible. Further, we would like to thank all professionals from Bartiméus and Royal Dutch Visio, and in particular Birgit van Iddekinge for her large contribution in developing the reading trainings. We would also like to thank students from the University of Groningen for their contribution in the data collection. Lastly, we would like to thank dr. Bart Melis-Dankers for his contribution to the development of this project and proofreading the manuscript.

