## Supplementary material for "Outcomes and Optimization of Two Reading Training Protocols for Individuals with Homonymous Visual Field Defects": S1 TIDieR checklists_1.pdf

### Supplementary file S1: The TIDieR (Template for Intervention Description and Replication) Checklist

Information to include when describing an intervention and the location of the information

| | | |
|---|---|---|
| **1. BRIEF NAME** | Vistra | Rotated Reading |
| **2. WHY** | The goals of the training are for the individual with HVFD to:<br>- experience less reading difficulties;<br>- learn to adjust the eye movements during reading in order to be less hindered by the HVFD;<br>- understand the influence of their HVFD on reading;<br>- be able to read different types of text in different circumstances. | The goals of the training are for the individual with HVFD to:<br>- experience less reading difficulties;<br>- be able to perceive bigger parts of words or sentence at once by reading rotated text;<br>- understand the influence of their HVFD on reading. |
| **3. WHAT: MATERIALS** | Vistra training software (contact:)<br>Handbook for clinician<br>Laptop<br>Computer monitor<br>Chinrest<br>Reading desk lamp<br>Book stand<br>Tablet, E-reader, Smartphone<br>Reading material: book, newspaper, magazine etc.<br>Magnifying glass<br>Homework material | Rotated Reading software (https://kennisoverzien.nl/interventies/rotated-reading-training)<br>Handbook for clinician<br>Laptop<br>Computer monitor<br>Reading desk lamp<br>Paper practice materials<br>Homework material |

| | | |
|---|---|---|
| **4. WHAT: PROCEDURES** | The seven themes within the training are: 1) insight and education on reading and HVFDs, 2) strategies to gain overview of different reading sources, 3) practice with adjusted eye movements, 4) for right-sided HVFDs: practice with adjusted eye movements during reading/keep reading; for left-sided HVFDs: practice with return sweep, 5) strategies to keep place/line in text, 6) practice with continued reading with even pace and 7) generalization of learned strategies to different reading objects and situations. To practice the adjusted eye movements, a software program is used with a fixation cross and stimuli in the blind hemifield with differing properties and presentation time. The participant starts practicing with single-figure stimuli, which builds up to words, sentences and eventually paragraphs. In between sessions, individuals with HVFDs are provided with daily homework assignments on paper which should take around 15 minutes to complete. After the last session, an evaluation takes place. Six weeks after the training, a follow-up evaluation by phone is performed by the occupational therapist. | Every session is made up of subgoals, comprised of a total of 10 subgoals. For session 1, these are: 1) determining the correct angle of rotation and 2) providing insight and education on reading and HVFDs. For session 2, these are: 3) gaining overview of rotated text, and 4) reading rotated words of maximal 7 letters. For session 3, the subgoals are 5) reading compound words of maximal 12 letters and 6) reading rotated sentences. In session 4, the subgoals are: 7) reading short rotated texts and 8) reading rotated meaningful texts (e.g. personal book of participant). Lastly, in session 5 the subgoals are 9) evaluation of the progress made and 10) discussion of how to implement the learned strategies in daily life. Practice assignments within the training are both on paper and computer. Further, the individual with a HVFD is asked to make daily homework assignments of 15 minutes (also both paper and digital). Six weeks after the training, a follow-up evaluation by phone is performed by the occupational therapist. |
| **5. WHO PROVIDED** | Occupational therapist with experience in HVFDs and followed specific Vistra training. | Occupational therapist with experience in HVFDs and followed specific Rotated Reading training. |
| **6. HOW** | Individual training sessions led by trained occupational therapist. | Individual training sessions led by trained occupational therapist. |
| **7. WHERE** | On-site at outpatient rehabilitation centre. | On-site at outpatient rehabilitation centre. |
| **8. WHEN and HOW MUCH** | According to protocol; around 8-10 weekly sessions of 60-90 minutes. | According to protocol: 5 weekly sessions of 60 minutes. |

| | | |
|---|---|---|
| **9. TAILORING** | The pace of the training (and thus amount of sessions) is determined by the speed of progress of the individual with a HVFD. The minimum amount of sessions is 6. The individual with a HVFD has the opportunity to practice with own reading materials as part of the homework assignments for sessions 4-6. | The pace of the training (and thus amount of sessions) is determined by the speed of progress of the individual with a HVFD. The minimum amount of sessions is 5. The individual with a HVFD has the opportunity to practice with own reading materials in session 4. |
| **10. MODIFICATIONS** | No modifications were made to the intervention protocol during the study. | No modifications were made to the intervention protocol during the study. |
| **11. HOW WELL** | Standardized session report forms were filled in by the occupational therapist. This form included for every session: an overview of the themes covered, if there was deviated from the protocol, whether the participant had made the homework and the concentration/fatigue levels of the participant. An overview of the filled in session report forms can be found on Supplementary file S2. | Standardized session report forms were filled in by the occupational therapist. This form included for every session: an overview of the subgoals covered, if there was deviated from the protocol, whether the participant had made the homework and the concentration/fatigue levels of the participant. An overview of the filled in session report forms can be found on Supplementary file S2. |
| **12. ACTUAL** | See session reports in Supplementary file S2. | See session reports in Supplementary file S2. |
| *Note.* HVFD = Homonymous Visual Field Defect. | | |
