## Supplementary material for "Outcomes and Optimization of Two Reading Training Protocols for Individuals with Homonymous Visual Field Defects": S2 Custom test.pdf

**Supplementary file S2: Example of the Custom Test (version right-sided HVFD for right eye)**

Figure without HVFD:

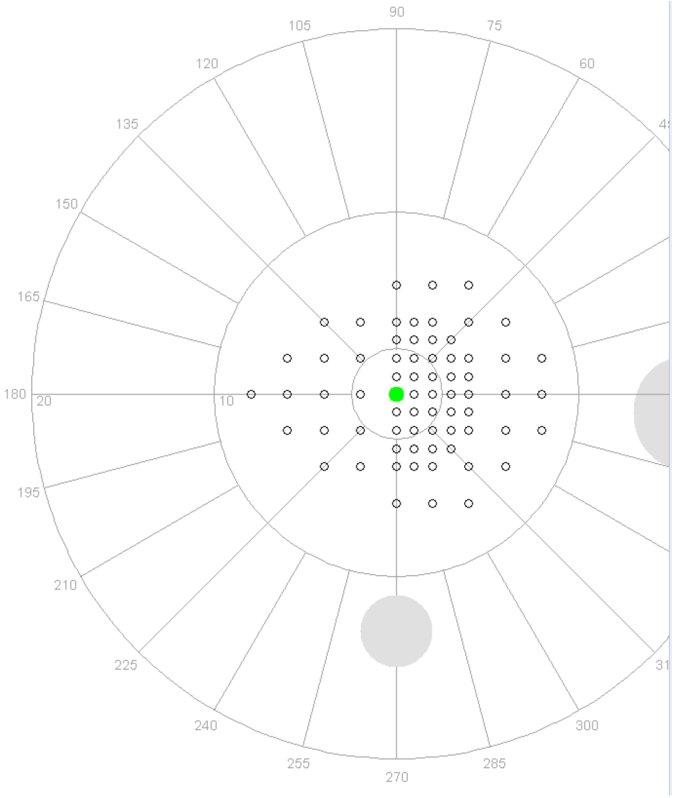


Example of output of participant with right-sided HVFD with approximately 1 degree macular sparing:

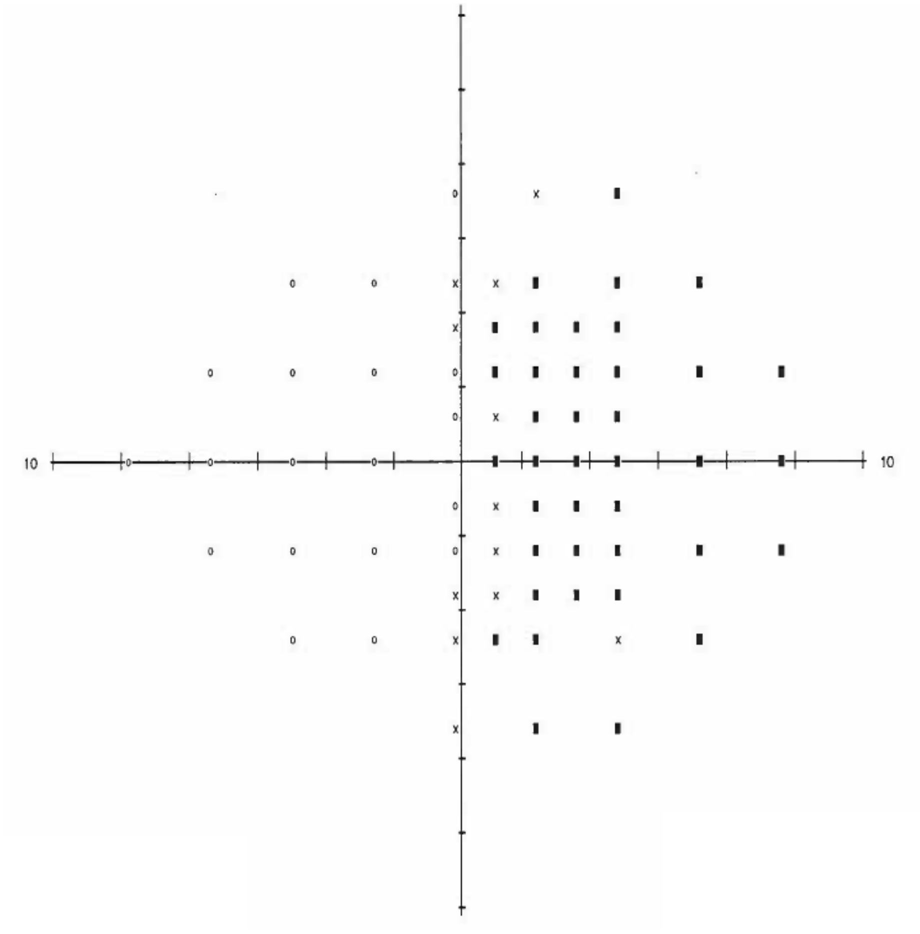
