## Supplementary material for "Outcomes and Optimization of Two Reading Training Protocols for Individuals with Homonymous Visual Field Defects": S3 Topic guide English.pdf

**Supplementary file S3: Topic Guides (translated)**

Please contact the authors for the original Dutch topic guide:

Translated Topic Guide

**Were your expectations about the training met?**

**Are you satisfied with the outcomes of the training?**

**What are the three most important things that the reading training has provided you with?**
1.
2.
3.

**How did you notice this?**

**In the pre-training assessment, you were asked about reading skills you hoped to improve with the reading training. Could you indicate whether there has been any improvement in those skills? At that time, you mentioned (fill in from HRQ):**
1
2
3

**In the pre-training assessment, you were asked about reading objects you hoped to readin better after the reading training. Could you indicate whether there has been any improvement in those objects? At that time, you mentioned (fill in from HRQ)::**
1
2
3

**How often do situations occur in daily life where you can apply what you have learned?**
(a) Very often
(b) Often
(c) Rarely
(d) Never

**When situations arise where you can apply what you have learned, how often do you apply what you have learned?**
(a) Very often
(b) Often
(c) Rarely
(d) Never

**What is a reason for you to apply what you have learned?**

**What is a reason for you not to apply what you have learned?**

**What is needed to continue using what you have learned?**

**Would you recommend the training to people in your situation?**
(a) Yes; **why?**
**(b)** No; **why not?**

**What did you experience as positive about the training?**

**Were there things you felt were missing in the training?**

**What rating, from 1 to 10, would you give for the entire training?**

**Is there anything else you would like to say about the training?**
