## Supplementary material for "Outcomes and Optimization of Two Reading Training Protocols for Individuals with Homonymous Visual Field Defects": S4 Comparisons effectiveness.pdf

**Supplementary file S4: Comparisons of Effectiveness**

| Outcome measure | Effect | Group(s) | Test | Analyses of effect |
|---|---|---|---|---|
| Reading speed (wpm) | Time | Vistra | rm | T1-T2-T3: F(1.75,7.00)=6.18, ***p*=.031**, $\eta^2_p$=.607, 1-β=1 |
| | | | t | T1-T2: t(4)=-3.96, ***p*=.017**, *d*=-1.772, 1-β=.84 |
| | | | t | T2-T3: t(4)=1.10, *p*=.335, *d*=.490 |
| | | | t | T1-T3: t(4)=-2.05, *p*=.110, *d*=-.916 |
| | | Rotated Reading | rm | T1-T2-T3: F(1.22,4.87)=.51, *p*=.554, $\eta^2_p$=.113 |
| | | Control | t | T1-T2: t(9)=-1.02, *p*=.333, *d*=.179 |
| | Group | Vistra vs Control | rm | T1-T2: F(1,13)=11.37, ***p*=.005**, $\eta^2_p$=.467, 1-β=1 |
| | | | t | T1: t(13)=-4.05, ***p*<.001**, *d*=-2.216, 1-β=.96 |
| | | | t | T2: t(13)=-2.481, ***p*=.028**, *d*=-1.359, 1-β=.63 |
| | | Rotated Reading vs Control | rm | T1-T2: F(1,13)=1.95, *p*=.186, $\eta^2_p$=.130 |
| | Time*Group | Vistra vs Control | rm | T1-T2: F(1,13)=4.86, ***p*=.046**, $\eta^2_p$=.272, 1-β=1 |
| | | Rotated Reading vs Control | rm | T1-T2: F(1,13)=.20, *p*=.662, $\eta^2_p$=.015 |
| Reading speed (wpm) silently read | Time | Vistra | t | T1-T2: t(4)=-1.43, *p*=.226, *d*=.640 |
| | | Rotated Reading | t | T1-T2: t(4)=-.76, *p*=.488, *d*=.341 |
| | | Control | t | T1-T2: t(4)=-.19, *p*=.857, *d*=.059 |
| | Group | Vistra vs Control | rm | T1-T2: F(1,13)=3.68, *p*=.077, $\eta^2_p$=.220 |
| | | Rotated Reading vs Control | rm | T1-T2: F(1,13)=2.15, *p*=.166, $\eta^2_p$=.142 |
| | Time*Group | Vistra vs Control | rm | T1-T2: F(1,13)=.276, *p*=.608, $\eta^2_p$=.021 |
| | | Rotated Reading vs Control | rm | T1-T2: F(1,13)=.007, *p*=.934, $\eta^2_p$=.001 |
| Reading efficacy | Time | Vistra | f | T0-T1-T2-T3: $\chi^2$(3)=11.13, ***p*=.011**, *W*=*.74*, 1-β=.81 |
| | | | w | T0-T1: Z=-2.06, ***p*=.039**, *r*=-.92, 1-β=1 |
| | | | w | T0-T2: Z=-1.84, *p*=.066. *r*=-.82 |
| | | | w | T0-T3: Z=-2.04, ***p*=.041**, *r*=-.91, 1-β=.87 |
| | | | w | T1-T2: Z=-1.47, *p*=.141, *r*=-.66 |
| | | | w | T1-T3: Z=-1.84, *p*=.066, *r*=-.82 |
| | | | w | T2-T3: Z=-.92, *p*=.357 *r*=-.41 |
| | | Rotated Reading | f | T0-T1-T2-T3: $\chi^2$(3)=10.64, ***p*=.014**, *W*=*.71*, 1-β=.79 |
| | | | w | T0-T1: Z=-2.06, ***p*=.039**, *r*=-.92, 1-β=1 |
| | | | w | T0-T2: Z=-1.84, *p*=.066 *r*=-.82 |
| | | | w | T0-T3: Z=-2.04, ***p*=.041**, *r*=-.91, 1-β=1 |
| | | | w | T1-T2: Z=-1.10, *p*=.273 *r*=-.49 |
| | | | w | T1-T3: Z=-1.63, *p*=.102 *r*=-.73 |
| | | | w | T2-T3: Z=-.58, *p*=.577 *r*=-.26 |
| | | Control | f | T1-T2: $\chi^2$(2)=11.49, ***p*=.003**, *W*=.52, 1-β=.87 |
| | | | w | T0-T1: Z=-2.53, ***p*=.011**, *r*=-.80, 1-β.99 |
| | | | w | T0-T2: Z=-2.57, ***p*=.010**, *r*=-.81, 1-β=.93 |
| | | | w | T1-T2: Z=-1.41 *p*=.160 *r*=-.45 |
| | Group | Vistra vs Control | m | T0: U=18, *p*=.362, $\eta^2$=.049 |
| | | | m | T1: U=14.5, *p*=.188, $\eta^2$=.110 |
| | | | m | T2: U=21, *p*=.616, $\eta^2$=.016 |
| | | Rotated Reading vs Control | m | T0: U=15, *p*=.181, $\eta^2$=.100 |
| | | | m | T1: U=24.5, *p*=.949, $\eta^2$=0 |
| | | | m | T2: U=19, *p*=.454, $\eta^2$=.036 |
| | Time*Group | Vistra vs Control | m | ΔT2-T1: U=17.5, *p*=.346, $\eta^2$=.056 |
| | | Rotated Reading vs Control | m | ΔT2-T1: U=20, *p*=.523, $\eta^2$=.025 |
| Reading attitude | Time | Vistra | f | T0-T1-T2-T3: $\chi^2$(3)=6.93, *p*=.074, *W*=.46 |
| | | Rotated Reading | f | T0-T1-T2-T3: $\chi^2$(3)=7.15, *p*=.067, *W*=.48 |
| | | Control | f | T1-T2: $\chi^2$(2)=6, *p*=.050, *W*=.30 |
| | Group | Vistra vs Control | m | T0: U=18.5, *p*=.384, $\eta^2$=.042 |
| | | | m | T1: U=18.5, *p*=.421, $\eta^2$=.042 |
| | | | m | T2: U=23.5, *p*=.825, $\eta^2$=.002 |
| | | Rotated Reading vs Control | m | T0: U=22.5, *p*=.690, $\eta^2$=.006 |
| | | | m | T1: U=16.5, *p*=.290, $\eta^2$=.072 |
| | | | m | T2: U=21, *p*=.619, $\eta^2$=.016 |
| | Time*Group | Vistra vs Control | m | ΔT2-T1: U=22, *p*=.708, $\eta^2$=.009 |
| | | Rotated Reading vs Control | m | ΔT2-T1: U=19, *p*=.455, $\eta^2$=.036 |
| Reading skills | Time | Vistra | f | T0-T1-T2-T3: $\chi^2$(3)=14.02, ***p*=.003**, *W*=.93, 1-β=.90 |
| | | | w | T0-T1: Z=-2.03, ***p*=.042**, *r*=-.91, 1-β=1 |
| | | | w | T0-T2: Z=-2.02, ***p*=.043**, *r*=-.90, 1-β=.94 |
| | | | w | T0-T3: Z=-2.03, ***p*=.042**, *r*=-.91, 1-β=.96 |
| | | | w | T1-T2: Z=-2.03, ***p*=.042**, *r*=-.91, 1-β=.99 |
| | | | w | T1-T3: Z=-2.03, ***p*=.042**, *r*=-.91, 1-β=1 |
| | | | w | T2-T3: Z=-.88, *p*=.705, *r*=-.39 |

| | | | | |
|---|---|---|---|---|
| | | Rotated Reading | f | T0-T1-T2-T3: $\chi^2$(3)=13, ***p*=.005**, *W*=.87, 1-β=.87 |
| | | | w | T0-T1: Z=-2.02, ***p*=.043**, *r*=-.90, 1-β=1 |
| | | | w | T0-T2: Z=-2.02, ***p*=.043**, *r*=-.90, 1-β=.94 |
| | | | w | T0-T3: Z=-2.03, ***p*=.042**, *r*=-.91, 1-β=1 |
| | | | w | T1-T2: Z=-2.03, ***p*=.042**, *r*=-.91, 1-β=.73 |
| | | | w | T1-T3: Z=-2.02, ***p*=.043**, *r*=-.90, 1-β=.30 |
| | | | w | T2-T3: Z=-.54, *p*=.593, *r*=-.24 |
| | | Control | f | T1-T2: $\chi^2$(2)=12.81, ***p*=.002**, *W*=.64, 1-β=.90 |
| | | | w | T0-T1: Z=-2.67, ***p*=.008**, *r*=-.84, 1-β=1 |
| | | | w | T0-T2: Z=-2.09, ***p*=.036**, *r*=-.66, 1-β=.60 |
| | | | w | T1-T2: Z=-1.26 *p*=.207, *r*=-.40 |
| | Group | Vistra vs Control | m | T0: U=20, *p*=.525, $\eta^2$=.025 |
| | | | m | T1: U=9, ***p*=.049,** $\eta^2$=.256, 1-β=.55 |
| | | | m | T2: U=25.5, *p*=.951, $\eta^2$=0 |
| | | Rotated Reading vs Control | m | T0: U=18, *p*=.364, $\eta^2$=.049 |
| | | | m | T1: U=4, ***p*=.010**, $\eta^2$=.441, 1-β=.83 |
| | | | m | T2: U=22.5, *p*=.759, $\eta^2$=.006 |
| | Time*Group | Vistra vs Control | m | ΔT2-T1: U=12, *p*=.109, $\eta^2$=.169 |
| | | Rotated Reading vs Control | m | ΔT2-T1: U=11, *p*=.084, $\eta^2$=.196 |
| Reading objects | Time | Vistra | f | T0-T1-T2-T3: $\chi^2$(3)=14.36, ***p*=.002**, *W*=.96, 1-β=.90 |
| | | | w | T0-T1: Z=-2.02, ***p*=.043**, *r*=-.90, 1-β=1 |
| | | | w | T0-T2: Z=-2.02, ***p*=.043**, *r*=-.90, 1-β=.99 |
| | | | w | T0-T3: Z=-2.02, ***p*=.043**, *r*=-.90, 1-β=.99 |
| | | | w | T1-T2: Z=-2.02, ***p*=.043**, *r*=-.90, 1-β=.98 |
| | | | w | T1-T3: Z=-2.02, ***p*=.043**, *r*=-.90, 1-β=1 |
| | | | w | T2-T3: Z=-.45, *p*=.655, *r*=-.20 |
| | | Rotated Reading | f | T0-T1-T2-T3: $\chi^2$(3)=14.13, ***p*=.003**, *W*=.94, 1-β=.90 |
| | | | w | T0-T1: Z=-2.02, ***p*=.043**, *r*=-.90, 1-β=.99 |
| | | | w | T0-T2: Z=-2.02, ***p*=.043**, *r*=-.90, 1-β=.85 |
| | | | w | T0-T3: Z=-2.02, ***p*=.043**, *r*=-.90, 1-β=.90 |
| | | | w | T1-T2: Z=-1.83, *p*=.068, *r*=-.82 |
| | | | w | T1-T3: Z=-1.75, *p*=.080, *r*=-.78 |
| | | | w | T2-T3: Z=-.37, *p*=.715, *r*=-.17 |
| | | Control | f | T1-T2: $\chi^2$(2)=12.51, ***p*=.002**, *W*=1.25, 1-β=.90 |
| | | | w | T0-T1: Z=-2.67, ***p*=.008**, *r*=-.84, 1-β=.98 |
| | | | w | T0-T2: Z=-2.52, ***p*=.012**, *r*=-.80, 1-β=.93 |
| | | | w | T1-T2: Z=-.18 *p*=.859, *r*=-.06 |
| | Group | Vistra vs Control | m | T0: U=22, *p*=.679, $\eta^2$=.009 |
| | | | m | T1: U=14, *p*=.178, $\eta^2$=.121 |
| | | | m | T2: U=14.5, *p*=.194, $\eta^2$=.090 |
| | | Rotated Reading vs Control | m | T0: U=20.5, *p*=.511, $\eta^2$=.020 |
| | | | m | T1: U=8.5, ***p*=.043**, $\eta^2$=.272, 1-β=.53 |
| | | | m | T2: U=20, *p*=.540, $\eta^2$=.025 |
| | Time*Group | Vistra vs Control | m | ΔT2-T1: U=2, ***p*=.005**, $\eta^2$=.529, 1-β=.96 |
| | | Rotated Reading vs Control | m | ΔT2-T1: U=11, *p*=.086, $\eta^2$=.196 |
| Vision-related quality of life | Time | Vistra | f | T1-T2-T3: $\chi^2$(2)=3.6, *p*=.165, *W*=.36 |
| | | Rotated Reading | f | T1-T2-T3: $\chi^2$(2)=1.2, *p*=.549, *W*=.12 |
| | | Control | w | T1-T2: Z=-2.07, ***p*=.038**, *r*=-.65, 1-β=.57 |
| | Group | Vistra vs Control | m | T1: U=22, *p*=.947, $\eta^2$=.009 |
| | | | m | T2: U=19, *p*=.641, $\eta^2$=.036 |
| | | Rotated Reading vs Control | m | T1: U=24, *p*=.903, $\eta^2$=.001 |
| | | | m | T2: U=11, *p*=.086, $\eta^2$=.196 |
| | Time*Group | Vistra vs Control | m | ΔT2-T1: U=21, *p*=.624, $\eta^2$=.016 |
| | | Rotated Reading vs Control | m | ΔT2-T1: U=9, *p*=**.050**, $\eta^2$=.256, 1-β=.82 |

Notes. Bolded values are significant (*p<.05).* rm = repeated measures anova. Due to small sample sized, the sizes the Greenhouse-Geisser correction was applied for repeated measures anova. t= t-test. d= Cohen's d. f=Friedman test. T0=retrospectively reported about the time before the HVFD, T1=pre-intervention assessment, T2=post-intervention assessment, T3=3-month follow-up assessment. w=Wilcoxon Signed Rank test. W=Kendall's W effect size. r=effect size for Wilcoxon Signed Rank test. m: Mann-Whitney U test. Δ = difference score between two time points. 1-β=achieved statistical power, post hoc calculated with software G*Power.
