## Supplementary material for "Outcomes and Optimization of Two Reading Training Protocols for Individuals with Homonymous Visual Field Defects": S5 Descriptives HRQ and VFQ.pdf

**Supplementary file S5: Descriptive Statistics of the WPM-silent, HRQ and NEI-VFQ-25**

Descriptive Statistics of Words Per Minute Silent

| | WPM-silent[a] | | | HVFD |
|---|---|---|---|---|
| Participant | T1 | T2 | *T2-T1* | side |
| V mean | 108.12 | 127.04 | +18.92 | |
| V1 | 75.53 | 142.53 | +67.00 | Left |
| V2 | 79.92 | 108.07 | +28.15 | Right |
| V3 | 122.85 | 120.32 | -2.53 | Left |
| V4 | 113.29 | 114.83 | +1.54 | Right |
| V5 | 148.99 | 149.44 | +.45 | Left |
| R mean | 121.31 | 127.29 | +5.98 | |
| R1 | 195.83 | 174.23 | -21.60 | Left |
| R2 | 96.08 | 106.72 | +10.64 | Right |
| R3 | 86.85 | 95.15 | +8.31 | Right |
| R4 | 63.44 | 90.46 | +27.02 | Right |
| R5 | 164.33 | 169.87 | +5.54 | Left |
| C mean[b] | 164.79 | 168.36 | +3.58 | |

*Note.* All participants had a macular sparing of less than 5 degrees. WPM=Words Per Minute, HVFD=Homonymous Visual Field Defect, V=Vistra, R=Rotated Reading, C=Control.

a. WPM calculated based on sum of reading times of 5 fragments of the IReST, digitally presented.
b. N=10.

Descriptive Statistics of the Hemianopia Reading Questionnaire

| | HRQ-r Efficacy (score 1-5) | | | | | | HRQ-r Attitude (score 1-5) | | | | | | HVFD side |
|---|---|---|---|---|---|---|---|---|---|---|---|---|---|
| Participant | T0 | T1 | T2 | T3 | *T2-T1* | *T3-T2* | T1 | T2 | T2 | T3 | *T2-T1* | *T3-T2* | |
| V total | 4.7 | 1.9 | 2.7 | 3.1 | +.8 | +4 | 4.5 | 3.8 | 3.9 | 4.3 | +.1 | +.3 | |
| V1 | 5.0 | 2.0 | 2.0 | 2.0 | +.0 | +.0 | 4.7 | 4.0 | 3.0 | 4.7 | -1.0 | +1.7 | Left |
| V2 | 5.0 | 2.5 | 3.5 | 4.0 | +1.0 | +.5 | 5.0 | 4.7 | 5.0 | 5.0 | +.3 | +.0 | Right |
| V3 | 4.0 | 1.5 | 4.0 | 3.0 | +2.5 | -1.0 | 4.3 | 3.3 | 4.3 | 4.7 | 1.0 | +.3 | Left |
| V4 | 4.5 | 1.0 | 2.0 | 3.0 | +1.0 | +1.0 | 4.3 | 3.0 | 3.3 | 3.3 | +.3 | +.0 | Right |
| V5 | 5.0 | 2.5 | 2.0 | 3.5 | -.5 | +1.5 | 4.3 | 4.0 | 4.0 | 3.7 | +.0 | -0.3 | Left |
| R total | 4.8 | 2.5 | 3.1 | 3.3 | +.6 | +.2 | 4.7 | 4.5 | 4.3 | 4.5 | -.2 | +.2 | |
| R1 | 5.0 | 3.0 | 3.0 | 3.0 | +.0 | +.0 | 5.0 | 4.7 | 4.7 | 5.0 | +.0 | +.3 | Left |
| R2 | 5.0 | 2.5 | 3.0 | 3.5 | +.5 | +.5 | 5.0 | 5.0 | 4.7 | 4.7 | -0.3 | +.0 | Right |
| R3 | 5.0 | 3.0 | 2.0 | 3.0 | -1.0 | +1.0 | 5.0 | 5.0 | 5.0 | 5.0 | +.0 | +.0 | Right |
| R4 | 4.0 | 2.0 | 4.0 | 3.0 | +2.0 | -1.0 | 3.7 | 3.7 | 3.0 | 3.3 | -0.7 | +.3 | Right |
| R5 | 5.0 | 2.0 | 3.5 | 4.0 | +1.5 | +.5 | 5.0 | 4.3 | 4.3 | 4.7 | +.0 | +.5 | Left |
| C total | 4.3 | 2.6 | 2.8 | | +.3 | | | | | | +.0 | | |

| | HRQ Skills (score 1-4) | | | | | | HRQ Objects (score 1-4) | | | | | | HVFD side |
|---|---|---|---|---|---|---|---|---|---|---|---|---|---|
| Participant | T0 | T1 | T2 | T3 | *T2-T1* | *T3-T2* | T1 | T2 | T2 | T3 | *T2-T1* | *T3-T2* | |
| V total | 3.8 | 2.2 | 2.8 | 2.8 | +.7 | +.0 | 3.9 | 2.5 | 3.1 | 3.1 | +.6 | +.0 | |
| V1 | 3.9 | 1.9 | 2.4 | 2.3 | +.5 | -0.1 | 4.0 | 2.4 | 3.0 | 3.0 | +.6 | +.0 | Left |

| | | | | | | | | | | | | | |
|---|---|---|---|---|---|---|---|---|---|---|---|---|
| V2 | 4.0 | 2.6 | 3.5 | 3.5 | +.9 | +.0 | 4.0 | 2.8 | 3.5 | 3.4 | +.7 | -0.1 | Right |
| V3 | 3.5 | 2.0 | 2.9 | 2.8 | +.9 | -0.1 | 3.4 | 2.6 | 3.0 | 3.0 | +.5 | +.0 | Left |
| V4 | 3.8 | 2.0 | 2.8 | 2.6 | +.8 | -0.1 | 3.9 | 2.0 | 3.0 | 3.0 | 1.0 | +.0 | Right |
| V5 | 4.0 | 2.3 | 2.6 | 2.9 | +.4 | +.3 | 4.0 | 2.8 | 3.1 | 3.3 | +.4 | +.2 | Left |
| R total | 3.8 | 2.0 | 2.7 | 2.7 | +.7 | -.1 | 3.8 | 2.2 | 2.5 | 2.5 | +.3 | +.0 | |
| R1 | 3.7 | 2.1 | 2.6 | 2.5 | +.4 | -0.1 | 4.0 | 2.3 | 2.7 | 2.4 | +.4 | -0.2 | Left |
| R2 | 4.0 | 1.9 | 2.8 | 3.0 | +.9 | +.3 | 4.0 | 2.1 | 2.4 | 2.4 | +.3 | +.0 | Right |
| R3 | 4.0 | 2.1 | 2.3 | 2.3 | +.1 | +.0 | 4.0 | 1.9 | 1.9 | 1.9 | +.0 | +.0 | Right |
| R4 | 3.4 | 2.1 | 3.0 | 2.5 | +.9 | -0.5 | 3.1 | 2.5 | 2.9 | 2.6 | +.4 | -0.3 | Right |
| R5 | 4.0 | 1.7 | 3.0 | 3.0 | +1.3 | +.0 | 4.0 | 2.4 | 2.7 | 3.1 | +.3 | +.4 | Left |
| C total | 3.6 | 2.6 | 2.8 | | +.2 | | 3.7 | 2.8 | 2.8 | | +.0 | | |

*Note.* HRQ=Hemianopia Reading Questionnaire, HRQ-r=HRQ Relationship to reading subscale, HVFD=Homonymous Visual Field Defect.

Descriptive Statistics of the Visual Function Questionnaire

| | NEI-VFQ-25 | | | | | HVFD |
|---|---|---|---|---|---|---|
| Participant | T1 | T2 | T3 | *T2-T1* | *T3-T2* | side |
| V total | 64.79 | 72.20 | 74.09 | +7.41 | +1.89 | |
| V1 | 51.29 | 69.85 | 59.36 | +18.56 | -10.49 | Left |
| V2 | 72.54 | 87.25 | 89.13 | +14.71 | +1.88 | Right |
| V3 | 58.67 | 64.47 | 66.82 | +5.80 | +2.35 | Left |
| V4 | 75.79 | 68.64 | 75.27 | -7.15 | +6.63 | Right |
| V5 | 65.67 | 70.79 | 79.88 | +5.12 | +9.09 | Left |
| R total | 64.03 | 61.01 | 65.98 | -3.02 | +4.97 | |
| R1 | 59.63 | 54.42 | 59.13 | -5.21 | +4.71 | Left |
| R2 | 61.50 | 67.46 | 75.29 | +5.96 | +7.83 | Right |
| R3 | 67.04 | 56.71 | 63.79 | -10.33 | +7.08 | Right |
| R4 | 63.38 | 55.17 | 60.79 | -8.21 | +5.62 | Right |
| R5 | 68.58 | 71.29 | 70.88 | +2.71 | -0.41 | Left |
| C total | 63.47 | 69.23 | | +5.76 | | |

*Note.* NEI-VFQ-25=National Eye Institute Visual Function Questionnaire 25-item, HVFD=Homonymous Visual Field Defect.
